# The relationship between controllability, optimal testing resource allocation, and incubation-latent period mismatch as revealed by COVID-19

**DOI:** 10.1101/2022.11.06.22281984

**Authors:** Jeffery Demers, William F. Fagan, Sriya Potluri, Justin M. Calabrese

## Abstract

The severe shortfall in testing supplies during the initial COVID-19 outbreak and ensuing struggle to manage the pandemic have affirmed the critical importance of optimal supply-constrained resource allocation strategies for controlling novel disease epidemics. To address the challenge of constrained resource optimization for managing diseases with complications like pre- and asymptomatic transmission, we develop an integro partial differential equation compartmental disease model which incorporates realistic latent, incubation, and infectious period distributions along with limited testing supplies for identifying and quarantining infected individuals. Our model overcomes the limitations of typical ordinary differential equation compartmental models by decoupling symptom status from model compartments to allow a more realistic representation of symptom onset and presymptomatic transmission. To analyze the influence of these realistic features on disease controllability, we find optimal strategies for reducing total infection sizes that allocate limited testing resources between ‘clinical’ testing, which targets symptomatic individuals, and ‘non-clinical’ testing, which targets non-symptomatic individuals. We apply our model not only to the original, delta, and omicron COVID-19 variants, but also to generically parameterized disease systems with varying mismatches between latent and incubation period distributions, which permit varying degrees of presymptomatic transmission or symptom onset before infectiousness. We find that factors that decrease controllability generally call for reduced levels of non-clinical testing in optimal strategies, while the relationship between incubation-latent mismatch, controllability, and optimal strategies is complicated. In particular, though greater degrees of presymptomatic transmission reduce disease controllability, they may increase or decrease the role of non-clinical testing in optimal strategies depending on other disease factors like transmissibility and latent period length. Importantly, our model allows a spectrum of diseases to be compared within a consistent framework such that lessons learned from COVID-19 can be transferred to resource constrained scenarios in future emerging epidemics and analyzed for optimality.

## 1 Introduction

Since its declaration as a global pandemic in March 2020 (1), COVID-19 has caused over 600 million cumulative infections and 6 million deaths (2). This loss of life and productivity together with ubiquitous lockdowns and mobility restrictions have resulted in devastating socioeconomic consequences worldwide (3, 4). The particularly severe costs of lockdowns underscore the need for effective large scale test-trace-quarantine programs to combat emerging disease epidemics and save lives while keeping society open and functioning. Unfortunately, for an epidemic caused by a novel pathogen, testing supplies and health care infrastructure may be inadequate to meet demand as health agencies struggle to implement new techniques and technologies at population-wide scales. During the initial phases of the COVID-19 pandemic, for example, testing capacities fell well-short of the levels required to monitor populations and test all suspected cases, while processing delays limited the usefulness of the tests that were actually available (5, 6). Even now, 3 years after the emergence of COVID-19, testing supplies have repeatedly struggled to meet the demand during case surges associated with new COVID variants (6).

When COVID resources have fallen short of demand, experts and health agencies have recommended the prioritization of testing supplies to the most severely symptomatic and vulnerable patients typically found in clinical health care settings, rather than to non-clinical cases associated with mildly or asymptomatic individuals and population monitoring programs (6, 7, 8). This strategy seeks to maximize the utility of the few resources available by limiting the amount ‘wasted’ on individuals who are not infected, but in the process excludes the possibility of slowing disease spread by identifying and quarantining non-symptomatic infected individuals (9). Achieving an ideal balance between clinical and non-clinical resource allocation is especially pertinent for a disease like COVID-19, where undetected presymptomatic transmission caused by mismatched latent and incubation periods (10, 11) as well as potentially large numbers of undetected totally asymptomatic spreaders (12, 13, 14, 15) present significant barriers to controllability.

Given the complexity of real-world disease dynamics with features like symptom-based testing and pre- and asymptomatic transmission, simplified compartmental mathematical models can provide powerful tools for analyzing and optimizing control strategies in the face of resource limitations. However, overly simplistic models can yield erroneous conclusions regarding real-world control strategies, so one must carefully balance model simplicity against the complex realistic elements most relevant to the problem at hand. Conventional compartmental COVID-19 control models are typically based on systems of ordinary differential equations (ODE’s) (16, 17, 18, 19, 20, 21, 22, 23, 24, 25). While ODE disease models provide a level of mathematical tractability, they necessitate the coupling of symptom status to specific model compartments, and this structural constraint can result in unnatural or unrealistic representations of symptom onset and presymtomatic transmission with potential unintended consequences on model behavior and real-world interpretations. This limitation is especially problematic when modeling controls like clinical and non-clinical testing strategies that are directly tied to symptom status.

ODE models for COVID-19 have generally addressed symptom onset and presymptomatic transmission by one of two broad schemes, both of which have their own drawbacks. One class of models simply ignores the potential for presymptomatic transmission by having infected individuals transition from an exposed non-symptomatic non-infectious compartment to an infectious symptomatic compartment, often with an additional infection channel comprised of permanently asymptomatic infected individuals. Such models have been used to analyze testing, contact tracing, and quarantine control strategies (16, 17), particularly in the context of limited resource constraints (18), along with vaccination control (19) and non-pharmaceutical interventions like masking and social distancing (20, 21). Although useful as simple baseline examples, these models may overestimate the efficacy of symptom-based COVID-19 controls due to the absence of presymptomatic transmission. A second more complicated class of ODE models includes a presymptomatic compartment where individuals are infectious but not yet symptomatic before transitioning to an infectious symptomatic compartment (22, 23, 24, 25). This approach, while more realistic, presents challenges when interpreting results for real world decision making via comparisons to the simpler class of models. Specifically, when multiple infectious stages are incorporated into an ordinary differential equation model, the total infectious period is no longer exponentially distributed (as would occur for a single infectious stage), but is instead more similar to a gamma or Weibull distribution (26), and such non-exponential distributions have been shown to be more difficult to control with identification-isolation strategies as compared to exponential distributions (27). Thus, it is unclear to what degree differences in output between models with and without presymptomatic compartments are due to presymptomatic transmission itself or to the presence of non-exponential infectious periods, and this presents a barrier to interpreting model results for real-world decision making. Furthermore, when a totally asymptomatic infection channel is included, one must either add a fictitious presymptomatic compartment for this class or accept a major qualitative difference between the infectious period distributions of those who will never and those who will eventually show symptoms (exponential vs non-exponential). Either option may have important impacts on model behavior, yet to our knowledge, this issue may has not been explicitly addressed in the literature. Finally, the class of models with presymptomatic compartments can not be adapted to other diseases for which infectiousness peaks well after symptom onset (such as with the 2003 SARS coronavirus (28, 29)) without significant changes to model structure, and this complicates any comparative mathematical analysis of diseases which are fundamentally similar to one another aside from changes in latent-incubation period mismatch. This deficit may be particularly problematic for utilizing mathematical models to inform real-world control strategies for new emerging epidemics based on the lessons learned from COVID-19.

We address these shortcomings by developing a partial integro differential equation model which utilizes the age of infection to decouple symptom status from specific model compartments. Our model is general in its ability to incorporate any latent, incubation, and infectious period distribution, regardless of shape or relative timings of means, and is utilized to analyze the original, delta, and omicron variants as well as a spectrum of other generically parameterized diseases all under a single lens. We incorporate a testing and quarantine control strategy which uses testing resources to identify infected individuals and remove them from the population while accounting for factors like contact tracing, limited population accessibility, and biases towards test-positive results. Our control assumes a fixed maximum testing capacity that must be allocated between clinical testing targeted at symptomatic individuals and non-clinical testing targeted at pre- and asymptomatic individuals. We analyze how controllability and optimal allocation strategies for reducing total infection size behave as functions of resource availability, testing quality, and disease characteristics. In general, we find that most factors that reduce controllability also call for a smaller share of resources to be devoted to non-clinical testing in optimal strategies. However, although presymptomatic transmission is found to reduce controllability, whether or not it reduces the role of non-clinical testing in optimal strategies depends on a complicated relationship between latent-incubation offset, disease transmissibility, and latent period length. In particular, we find that diseases with presymptomatic transmission do not necessarily call for a increase in non-clinical testing resource allocation compared to diseases lacking presymptomatic transmission, despite the fact that intuition would deem non-clinical testing to be of greater importance due to its the potential to eliminate presymptomatic spreaders. Together, these results highlight how intuition for disease control strategies based on qualitative disease characteristics may fail and thus emphasize the need for mathematical modeling to prepare for and manage future epidemics.

## 2 Methods

We extend the previous ordinary differential equation (ODE) SEIR testing and quarantine model of Calabrese and Demers (18) to a system of integro-partial differential equations (IPDE) which explicitly incorporate the age of infection for infected classes. While our primary focus is COVID-19, including the original, delta, and omicron variants, our system is general in its ability to account for any set of latent, incubation, and infectiousness periods.

### 2.1 Model outline

#### 2.1.1 Uncontrolled transmission model

We assume a homogeneously mixed system of *N* total susceptible, exposed, infectious, and recovered individuals. Exposed and infectious classes are partitioned into those who will remain permanently asymptomatic throughout the course of the infection and those who will eventually become symptomatic at some point before recovery. Here, we take “asymptomatic” to mean genuinely exhibiting no symptoms or exhibiting symptoms so minor that one would not typically consider themselves ill or seek medical attention, and “symptomatic” to mean exhibiting visually identifiable symptoms with moderate to critical illness. Upon infection, initially susceptible individuals *S* will enter either the exposed permanently asymptomatic class *E*_*A*_ or the exposed eventually symptomatic class *E*_*Y*_ with probabilities *f*_*A*_ and *f*_*Y*_, respectively (throughout this paper, we use the abbreviations PA and ES to denote permanently asymptomatic and eventually symptomatic individuals). While in an exposed class, individuals are infected but not yet infectious and are thus unable to transmit the disease to others. To account for the possibility of non-exponential waiting times between infection sages, we continuously index exposed classes with the age of infection *x*. The number of individuals in the exposed PA class at time *t* who have been infected between *x* and *x* + *dx* units of time is denoted *e*_*A*_(*t, x*)*dx* such that the total number of exposed PA individuals *E*_*A*_(*t*) is given By 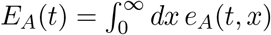. The age of infection-indexed exposed ES class *e*_*Y*_ (*t, x*) is defined analogously. PA and ES exposed individuals with infection age *x* transition to their respective infectious classes *a*(*t, x*) and *y*(*t, x*) at rate *ε*(*x*). Infectious individuals enter into the recovered class *R* where they are no longer infectious and are assumed to attain permanent immunity at age of infection dependent recovery rate *r*(*x*). Although there is some evidence suggesting that ES and PA individuals recover from COVID-19 at different rates, disease progression in asymptomatic individuals is poorly understood, and conflicting studies have shown faster, similar, and slower viral clearance rates in asymptomatic versus symptomatic cases (30, 31). For simplicity, we therefore take *r*(*x*) to be equivalent for PA and ES infections. Likewise, we are aware of only one study suggesting different transitions rates from the exposed to infectious class for PA versus ES individuals (10), but the corresponding average waiting times differ by only 5%, so we assume exposed to infectious transition rates to be independent of the eventual presence or lack of symptoms. The dynamical equations for the uncontrolled transmission model are as follows:

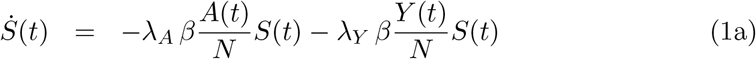

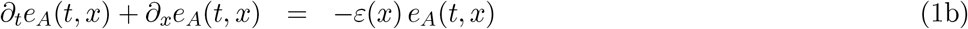

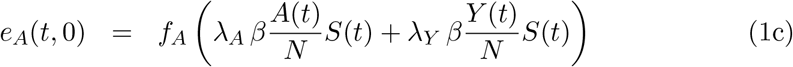

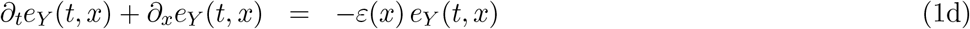

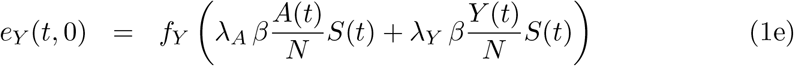

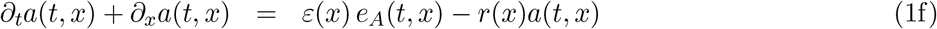

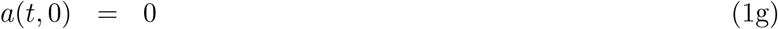

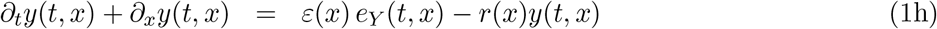

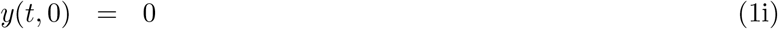

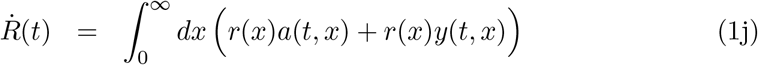

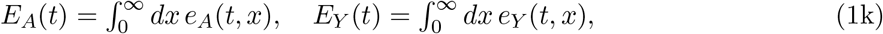

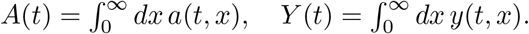

In the above equations, overdots denote ordinary derivatives with respect to time *t, β* denotes the average number of contacts made per unit time by an individual, *N* is the total population size, and *λ*_*A*_ and *λ*_*Y*_ denote the transmission probability per susceptible-infectious contact for the PA and ES classes, respectively (for simplicity, we assume transmission probability to be independent of the age of infection). The boundary terms *e*_*A*_(*t*, 0) and *e*_*Y*_ (*t*, 0) denote the rates of newly generated infections and thus represent to individuals with infection age *x* = 0. The boundary terms *a*(*t*, 0) and *y*(*t*, 0) are zero due to the fact that every infected individual will spend at least some time in an exposed class before becoming infectious and will therefore never enter an infectious class with an infection age *x* = 0. The integrals in Eq. (1k) relate the total number of individuals in an infected class to the corresponding distribution over the age of infection.

#### 2.1.2 Symptom onset

The model in Eq. (1) makes no assumptions regarding the onset of symptoms in infected individuals. This modeling choice is based on the fact that the natural dynamics of disease transmission (absent interventions or controls explicitly correlated with symptom status) depend fundamentally on infectious states rather than symptom states. In our IPDE model, symptoms are an incidental background state which need not be represented by separate pre- and post-symptom onset compartments for each infected compartment as would be required in an ordinary differential equation (ODE) model. We utilize the age of infection in the ES classes to the define the symptomatic population *X*_*S*_(*t*) by the following integral:

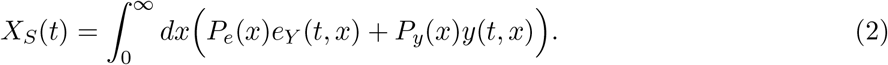

The functions *P*_*e*_(*x*) and *P*_*y*_(*x*) denote the respective probabilities for exposed and infectious individuals to show symptoms by infection day *x*. These probabilities likewise determine the nonsymptomatic infected population *X*_*N*_ (*t*) as follows:

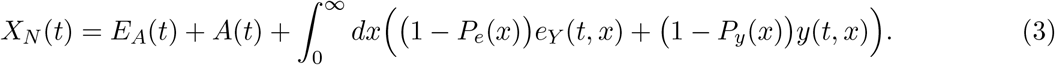

The non-symptomatic infected population is comprised of all PA infected individuals plus the ES infected individuals who are not yet showing symptoms.

In our simulations, we consider two assumptions for *P*_*e*_(*x*) and *P*_*y*_(*x*), “correlated symptoms” and “incubation symptoms:”

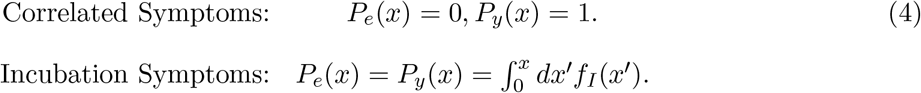

The correlated symptoms assumption defines symptom onset to occur in perfect correlation with infectiousness onset. Here, there is no possibility of pre-symptomatic transmission or pre-infectious symptom onset, and the symptomatic population is the entire ES infectious class. This assumption is equivalent to the symptom assumptions of our previous ODE testing and quarantine COVID-19 model in (18). The incubation symptoms assumptions defines *P*_*e*_(*x*) and *P*_*y*_(*x*) to be the cumulative distribution function of an incubation period distribution *f*_*I*_(*x*). Here, *f*_*I*_(*x*)*dx* denotes the probability for an ES individual to begin showing symptoms between age of infection *x* and *x* + *dx*. The flow diagram in Fig. 1 summarizes the progression of infectious and symptomatic states in the uncontrolled transmission model.

**Figure 1.**
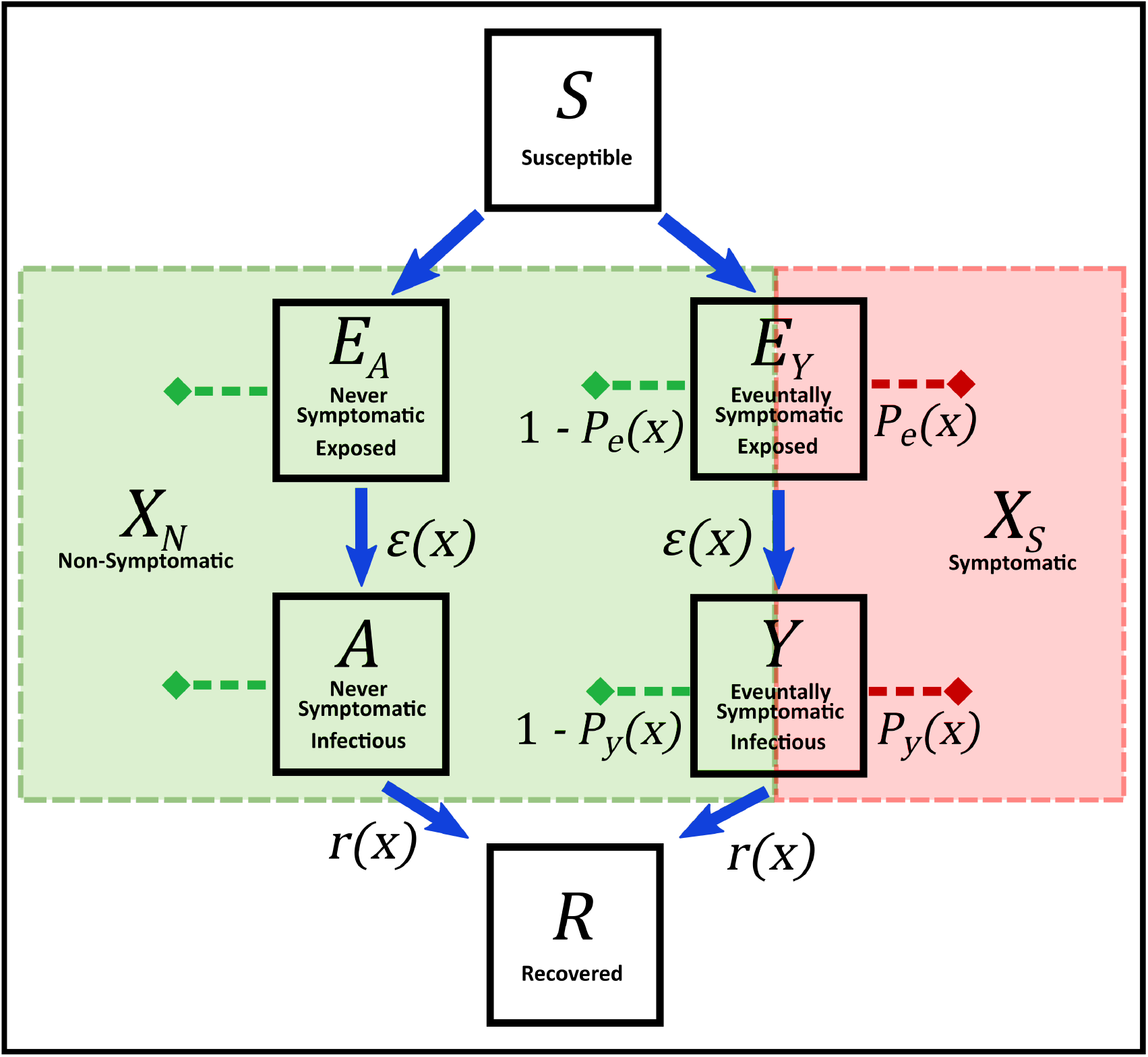
Schematic indicating the flow of infectious and symptomatic states in our uncontrolled transmission model. Upon infection, susceptible individuals *S* move into either the permanently asymptomatic (PA) exposed class *E*_*A*_ or the eventually symptomatic (ES) exposed class *E*_*Y*_ with an initial age of infection *x* = 0. As individuals linger in the exposed classes, their ages of infection increase, and transitions to the PA infectious class *A* and ES infectious class *Y* occur at infection age dependent transition rate *ε*(*x*) Infectious individuals then transition to the recovered class *R* at the infection-age-dependent transition rate *r*(*x*). Symptom status appears as background state indicated by the red and green boxes. For ES exposed and infectious individuals with infection age *x*, a fraction *P*_*e*_(*x*) and *P*_*y*_(*x*), respectively, will be in the symptomatic state *X*_*S*_, while the remaining fractions 1 − *P*_*e*_(*x*) and 1 − *P*_*y*_(*x*) will be in the non-symptomatic state *X*_*N*_. The PA classes *E*_*A*_ and *A* remain in the non-symptomatic state *X*_*N*_ until recovery.

#### 2.1.3 Testing and quarantine transmission model

We adapt the resource allocation testing and quarantine control framework from the ODE model of (18) to our IPDE disease model. Testing identifies and transfers infected individuals to a quarantine class *Q* where they remain isolated from contacts until recovery and are unable to generate new transmissions. We assume a finite testing resource represented by the testing capacity *C*, defined as the maximum per capita number of tests able to be administered and processed per day. A fraction *ρ* of the testing capacity is allocated to non-clinical testing for identifying non-symptomatic infected individuals, while the remaining fraction 1 *− ρ* is allocated to clinical testing for identifying symptomatic infected individuals. Clinical testing is conducted only on individuals showing sufficiently severe visually identifiable symptoms, while non-clinical testing is accessible to the general public. The actual rate at which tests are administered and processed for the two testing categories is dependent on the *testing demand*, i.e. on the number of people eligible for and actively seeking testing. Here, we provide the functional forms of test administration and processing rates for clinical and non-clinical testing and sketch the reasoning behind their formulation. Further details are elucidated in (18).

The average time required for a non-clinical test to be administered and processed for a single individual is denoted *τ*_*N*_ such that the average administration and processing rate is given by 1*/τ*_*N*_ :

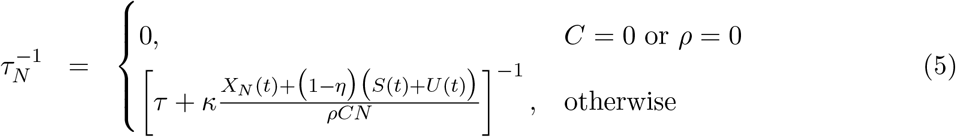

Non-clinical testing enters the disease model by transferring a fraction *κ* of the non-symptomatic infected population (either pre- or asymptomatic) to the quarantined class *Q* at rate 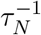. The case *C* = 0 corresponds to no testing capacity and thus no control, while the case *ρ* = 0 corresponds to all resources being devoted to clinical testing which reduces the non-clinical testing rate to zero. The quantity *τ* is the *intrinsic testing time*, defined as average time required for an individual get to a testing center, get tested, and receive results absent of delays or backlogs due to other patents. We take this value to be equal to one day. The testing demand for non-clinical testing is defined as a fraction *κ* the non-symptomatic population *X*_*N*_ (*t*) in Eq. (3) as well as a fraction *κ*(1 *− η*) of the uninfected population *S*(*t*) + *U* (*t*). Here, *U* (*t*) represents individuals who were previously infected and subsequently recovered without being tested and are thus unaware that they have immunity (we assume recovered individuals who were tested and quarantined know that they have immunity and therefore exclude themselves from future testing). The *accessibility parameter κ ∈* (0, 1] represents the fraction of the total population open to and compliant with non-clinical testing, and the *concentration parameter η ∈* [0, 1) represents the degree to which testing is focused on or biased towards infected individuals. A purely random population monitoring program which reaches all members of the population corresponds to *κ* = 1 and *η* = 0. Any factor that influences a given test such that it will be more likely applied to an infected individual as compared to purely random sampling will increase *η*. For example, those who suspect a recent disease exposure may be both more likely to seek testing and more likely to test positive compared a randomly selected individual, and this influence will focus testing towards the *X*_*N*_ population and away from the *S* +*U* population. Likewise, a highly effective contact tracing program may result in a relatively large value of *η*, but there may be many individuals unwilling to participate in government or health agency efforts, so the corresponding value of *κ* may be relatively small. Generally, the differing behavioral characteristics of individuals and differing testing policies of local governments unique to differing regions will correspond to a variety of concentration and accessibility levels, so we will analyze model results for a wide array of *κ* and *η* values. The analysis in (18) shows that *η* can be estimated from data comparing test-positive to disease prevalence rates, and that *η* = 0.95 is an upper bound on optimistically achievable values.

The average time required for a clinical test to be administered and processed for a single symptomatic individual is denoted *τ*_*C*_ such that the average administration and processing rate is given by 1*/τ*_*C*_:

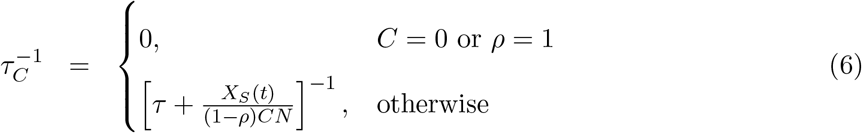

Clinical testing enters the disease model by transferring symptomatic individuals to the quarantine class *Q* at rate 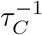. The case *C* = 0 corresponds to no control, while the case *ρ* = 1 corresponds to all resources being devoted to non-clinical testing which reduces the clinical testing rate to zero.

In contrast to non-clinical testing, clinical testing is assumed to have full concentration on and full accessibility to the target infected population, meaning that the testing demand is comprised only of the symptomatic population *X*_*S*_ and includes no uninfected individuals. We base this on the assumption that all individuals in the ES classes who are currently showing symptoms are all ill enough to seek medical attentions (full advisability), and that due to the presence of strong visually identifiable symptoms and the lack of secondary diseases in our model, there is no chance of using a clinically allocated test on an uninfected individual (full concentration). In this sense, clinical testing acts as a verification tool, while non-clinical testing acts as an identification tool.

For both testing types, when testing demand is very low, tests are administered and processed at maximum rates 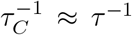 and 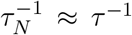 per individual, and the total number of clinical tests conducted per day grows linearly with the size of the symptomatic population. As demand increases, supply limitations and patient backlogs cause 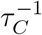 and 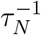 to decrease towards zero, and the total number of clinical and non-clinical tests conducted per day saturates to (1 − *ρ*)*CN* and *ρCN*, respectively, as testing demand approaches infinity. The full model equations with testing and quarantine control are given in Eq. (7) and a corresponding flow diagram is given in Fig. 2. Table 1 summarizes the definitions of all control-related parameters.

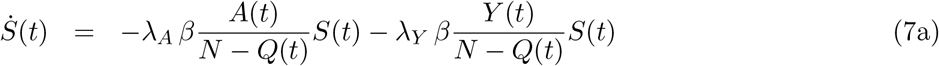

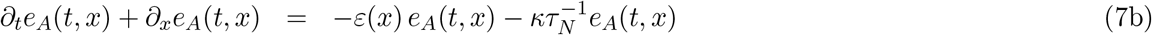

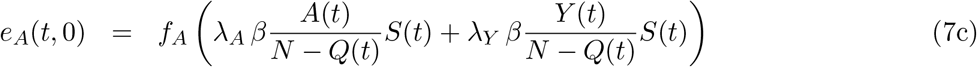

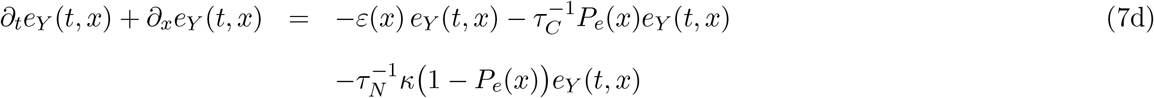

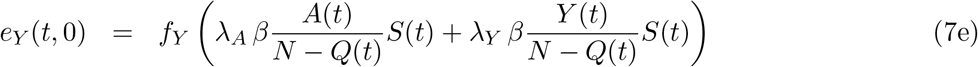

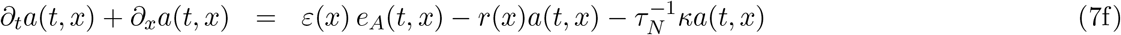

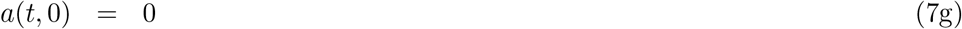

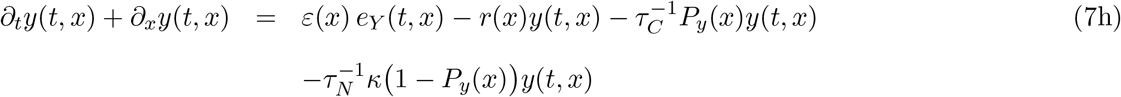

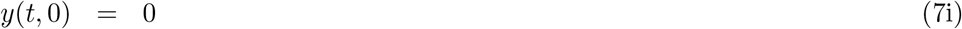

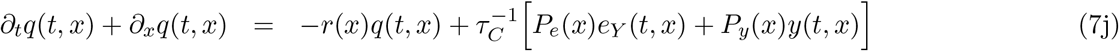

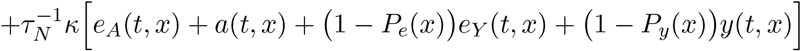

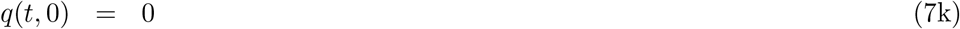

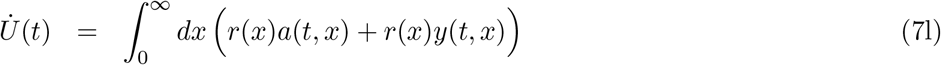

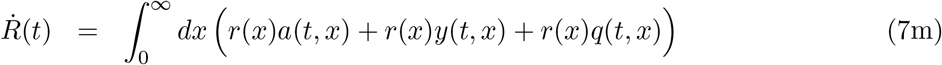

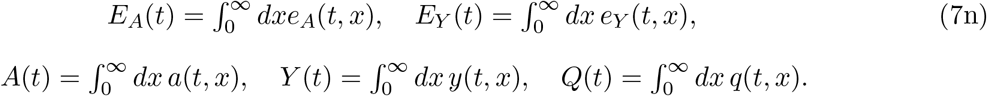

**Figure 2.**
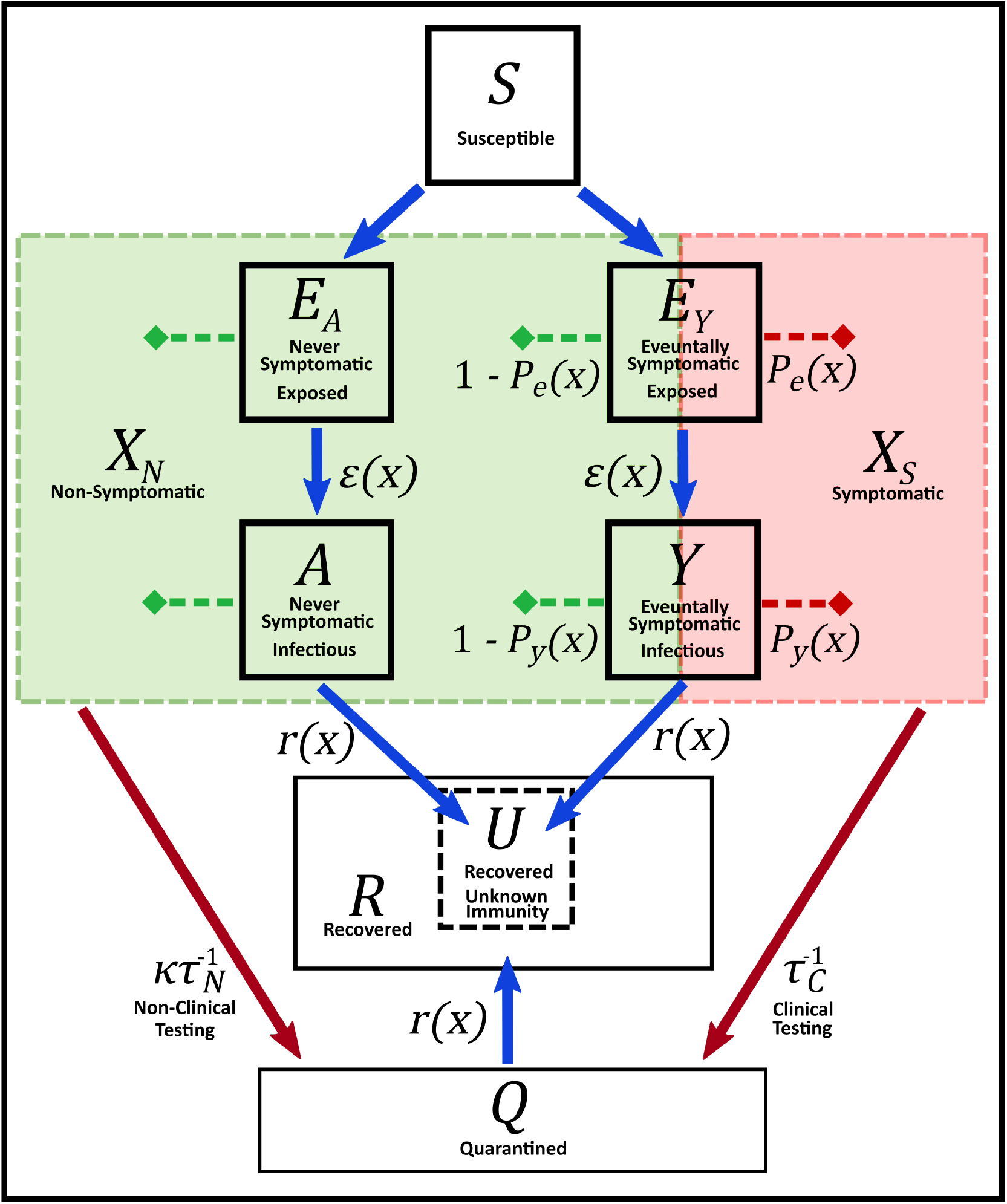
Flow diagram for the testing and quarantine control model in Eq. (7). The symptomatic population *X*_*S*_ and fraction *κ* of the non-symptomatic population *X*_*N*_ transition to the quarantine class *Q* by clinical and non-clinical testing, respectively, at rates 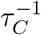 and 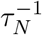. Quarantined individuals transition to the recovered class *R* at age of infection dependent rate *r*(*x*). The subset *U* of the recovered class represents individuals who were previously infected and subsequently recovered without testing and quarantine and are thus unaware that they have obtained immunity.

**Table 1:** Testing and quarantine control parameter definitions

| Parameter | Name | Meaning |
| --- | --- | --- |
| $C$ | Testing capacity | Maximum number of tests able to be administered and processed per day per capita |
| $\tau$ | Testing time | Average amount of time required for an individual be tested (including procrastination, travel time, processing time, etc.) absent of backlogs or delays due to other patients |
| $\rho$ | Strategy parameter | Fraction of testing capacity devoted to non-clinical testing |
| $\eta$ | Concentration parameter | $(1 - \eta)$ = Degree to which non-clinical testing resources are utilized on uninfected individuals |
| $\kappa$ | Accessibility parameter | Fraction of eligible population open to and compliant with non-clinical testing |

#### 2.1.4 Transition rates

The transition rates *ε*(*x*) and *r*(*x*) are determined by probability distributions for infection ages at which transitions between disease states occur. Specifically, letting *f*_*ε*_(*x*) denote the probability density for an infected individual to transition from exposed to infectious at infection age *x*, and letting *f*_*r*_(*x*) denote the probability density for an infectious individual to transition to recovered at infection age *x*, the corresponding transition rates are defined as follows:

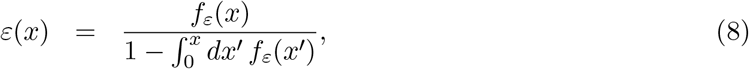

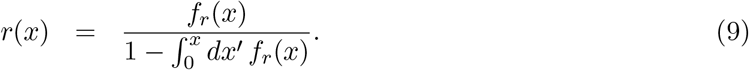

These transition rates represent conditional probability densities for an individual to transition to the next disease stage on infection day *x* given that they are still in the preceding disease stage up to day *x*. The probability densities for the infectious period duration and total infection duration, denoted *f*_*inf*_ (*x*) and *f*_*tot*_(*x*), respectively, can be written in terms of *f*_*ε*_(*x*) and *f*_*r*_(*x*) as follows:

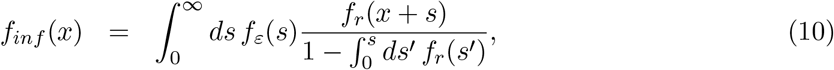

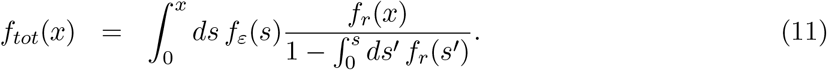

To clarify, *f*_*tot*_(*x*)*dx* represents the probability for an newly infected individual to remain infected for a total duration between *x* and *x* + *dx* days, *f*_*inf*_ (*x*)*dx* represents the probability for that individual to be infectious (i.e. contagious) for a total duration between *x* and *x* + *dx* days over the course of their infection, and *f*_*r*_(*x*)*dx* represents the probability for that individual to recover between infection days *x* and *x* + *dx* given that they have already entered into an infectious state. If *f*_*r*_(*x*) is assumed to be an exponential distribution with mean 1*/r, r*(*x*) reduces to a constant *r*(*x*) = *r* while *f*_*inf*_ (*x*) becomes equivalent to *f*_*r*_(*x*). Likewise, if *f*_*ε*_(*x*) is exponential with mean 1*/ε, ε*(*x*) reduces to a constant *ε*(*x*) = *ε*. If both *f*_*r*_(*x*) and *f*_*ε*_(*x*) are exponential, *f*_*tot*_(*x*) becomes a two parameter hypoexponential distribution with parameters *ε* and *r*. The exponential distribution assumptions taken together with the correlated symptoms assumption in Sec. 2.1.2 reduce the IPDE testing and quarantine model in Eq. (7) to the ODE testing and quarantine model in (18).

### 2.2 Distribution and parameter values

#### 2.2.1 COVID-19 variants

The distributions from which the transition rates *ε*(*x*) and *r*(*x*) are calculated, as well as the incubation period distribution, are based on epidemiological data for the original, delta, and omicron COVID-19 variants. The incubation period distribution *f*_*I*_(*x*) is a widely studied quantity for the early strains of COVID-19 which has been variously fit to log-normal, gamma, or Weibull distributions (32, 33, 34, 35, 36, 37). The latent period distribution *f*_*ε*_(*x*) and post-infectiousness onset recovery day distribution *f*_*r*_(*x*) are comparatively less well studied. For the original COVID-19 strain and delta variant, we utilize the results of Xin et al. (10) and Kang et al. (11), respectively, which estimate both *f*_*ε*_ and *f*_*I*_ to be gamma distributions. We are aware of only one data-based study estimating the incubation period distribution for the omicron variant, Tanaka et al. (38), which fits to a log-normal distribution with median 2.8 days. Other investigations estimating medians but not distributions have likewise obtained results approximately equal to 3 days (39, 40). To maintain consistency with the original and delta variants, we assume *f*_*I*_ for omicron to be gamma distributed with mean and variance comparable to that of (38). We are unaware of any existing estimates for the omicron variant’s latent period distribution, so we assume a gamma distribution with mean 1.5 days shorter than that of the incubation period. For all variants, we are unaware of any studies that directly estimate the post-infectiousness onset recovery day distribution *f*_*r*_(*x*). However, viral culturing studies have consistently found live viral isolation to cease between 9 and 10 days post-symptom onset (30, 31, 41). We therefore assume a gamma distribution for *f*_*r*_(*x*) with mean ⟨*f*_*r*_⟩ = ⟨*f*_*I*_ ⟩ + 9.5 days for each variant, and we assume a scale parameter of 0.25 days to give tight distributions with relatively small interquartile ranges as indicted in (41).

To assess the impact of the PDE elements of our model relative to the corresponding ODE model, we will compare simulation results under exponential and gamma distribution assumptions for *f*_*ε*_ and *f*_*r*_. Likewise, to assess the impact of pre-symptomatic transmission on disease controllability, we will compare model results between the correlated symptoms and incubation symptoms assumptions in Eq. (4). The means ⟨*f*_*ε*_ ⟩ are taken to be equivalent under the exponential and gamma assumptions, while we set ⟨*f*_*r*_ ⟩ = ⟨*f*_*I*_ ⟩ *−* ⟨*f*_*ε*_ ⟩ + 9.5 days under the exponential assumption as compared to ⟨*f*_*r*_ ⟩ = ⟨*f*_*I*_ ⟩ + 9.5 days under the gamma assumption. This difference ensures that the mean infectious period duration ⟨*f*_*inf*_ ⟩ and mean total infection time ⟨*f*_*tot*_ ⟩ are essentially unchanged by the different assumptions on *f*_*r*_. The means and standard deviations of *f*_*ε*_, *f*_*r*_, and *f*_*I*_, as well as that of the resultant distributions *f*_*inf*_ and *f*_*tot*_, are given in Tables 2, 3, and 4 for the original, delta, and omicron COVID variants, respectively. Plots depicting the shapes of *f*_*ε*_, *f*_*r*_, and *f*_*I*_ for the three variants are given in Fig. 3. For all variants and distribution assumptions, the average infectious period is approximately 11 days while the average latent period is approximately 1.5 days shorter than the average incubation period, thus indicating an average 1.5 day presymptomatic transmission window under the incubation symptoms assumption. The mean total infection duration decreases from the original to delta to omicron variant.

**Figure 3.**
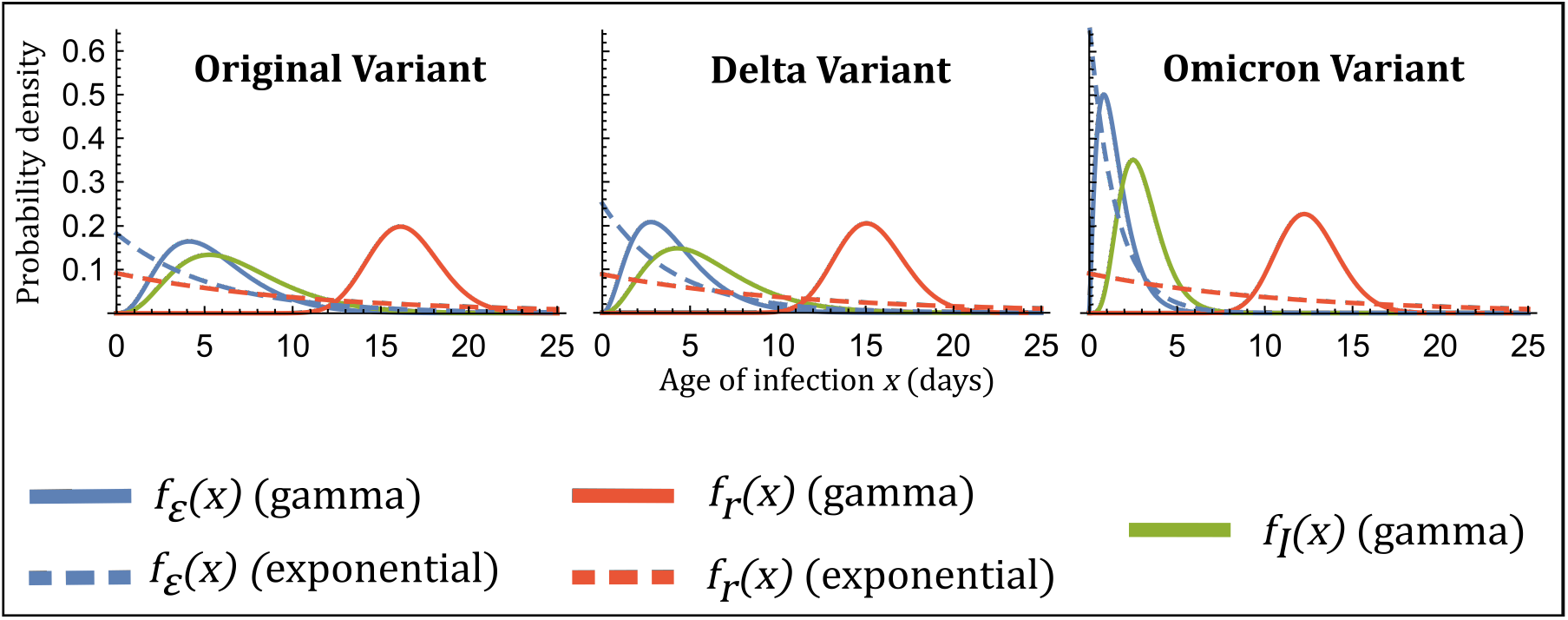
Latent period distributions *f*_*ε*_(*x*), incubation period distributions *f*_*I*_(*x*), and post-infectiousness onset recovery day distributions *f*_*r*_(*x*) as functions of the age of infection *x* for the original, delta, and omicron variants of COVID-19. Model simulations will consider combinations of exponential (dotted lines) and gamma (solid lines) distribution assumptions for *f*_*ε*_ and *f*_*r*_, as well as an incubation period symptom onset assumption using a gamma distribution for *f*_*I*_ or a correlated symptom assumption where symptom onset coincides exactly with the onset of infectiousness.

**Table 2:** Original variant distributions based on Refs. (30, 31, 10, 41). Simulations assume either exponential or gamma distributions for *f*_*ε*_ and *f*_*r*_ with means ⟨ ⟩ and standard deviations *σ* as indicated in units of days. Means and standard deviations for the resulting infectious period distribution *f*_*inf*_ and total infection duration distribution *f*_*tot*_ are likewise indicated. The incubation symptoms assumption utilizes a gamma distributed incubation period length *f*_*I*_ with indicated mean and standard deviation in units of days.

| Original Variant | $f_\varepsilon$ Gamma Distribution | | $f_\varepsilon$ Exponential Distribution | |
| --- | --- | --- | --- | --- |
| $f_r$ Gamma Distribution | $\langle f_\varepsilon \rangle = 5.48$ | $\sigma_\varepsilon = 2.72$ | $\langle f_\varepsilon \rangle = 5.48$ | $\sigma_\varepsilon = 5.48$ |
| | $\langle f_r \rangle = 16.38$ | $\sigma_r = 2.02$ | $\langle f_r \rangle = 16.38$ | $\sigma_r = 2.02$ |
| | $\langle f_{inf} \rangle = 10.92$ | $\sigma_{inf} = 3.34$ | $\langle f_{inf} \rangle = 11.26$ | $\sigma_{inf} = 4.81$ |
| | $\langle f_{tot} \rangle = 16.40$ | $\sigma_{tot} = 2.02$ | $\langle f_{tot} \rangle = 16.74$ | $\sigma_{tot} = 2.69$ |
| $f_r$ Exponential Distribution | $\langle f_\varepsilon \rangle = 5.48$ | $\sigma_\varepsilon = 2.72$ | $\langle f_\varepsilon \rangle = 5.48$ | $\sigma_\varepsilon = 5.48$ |
| | $\langle f_r \rangle = 10.90$ | $\sigma_r = 10.90$ | $\langle f_r \rangle = 10.90$ | $\sigma_r = 10.90$ |
| | $\langle f_{inf} \rangle = 10.90$ | $\sigma_{inf} = 10.90$ | $\langle f_{inf} \rangle = 10.90$ | $\sigma_{inf} = 10.90$ |
| | $\langle f_{tot} \rangle = 16.38$ | $\sigma_{tot} = 11.23$ | $\langle f_{tot} \rangle = 16.38$ | $\sigma_{tot} = 12.20$ |
| $f_I$ Gamma Distribution: $\langle f_I \rangle = 6.88$ $\sigma_I = 3.32$ | | | | |

**Table 3:** Delta variant distributions based on Refs. (30, 31, 11, 41). Meanings of quantities are as described in Table 2.

| Delta Variant | $f_\varepsilon$ Gamma Distribution | $f_\varepsilon$ Exponential Distribution |
| --- | --- | --- |
| $f_r$ Gamma Distribution | $\langle f_\varepsilon \rangle = 4.00$ $\sigma_\varepsilon = 2.22$<br>$\langle f_r \rangle = 15.30$ $\sigma_r = 1.96$<br>$\langle f_{inf} \rangle = 11.30$ $\sigma_{inf} = 2.94$<br>$\langle f_{tot} \rangle = 15.30$ $\sigma_{tot} = 1.96$ | $\langle f_\varepsilon \rangle = 4.00$ $\sigma_\varepsilon = 4.00$<br>$\langle f_r \rangle = 15.30$ $\sigma_r = 1.96$<br>$\langle f_{inf} \rangle = 11.43$ $\sigma_{inf} = 4.02$<br>$\langle f_{tot} \rangle = 15.43$ $\sigma_{tot} = 2.14$ |
| $f_r$ Exponential Distribution | $\langle f_\varepsilon \rangle = 4.00$ $\sigma_\varepsilon = 2.22$<br>$\langle f_r \rangle = 11.30$ $\sigma_r = 11.30$<br>$\langle f_{inf} \rangle = 11.30$ $\sigma_{inf} = 11.30$<br>$\langle f_{tot} \rangle = 15.30$ $\sigma_{tot} = 11.51$ | $\langle f_\varepsilon \rangle = 4.00$ $\sigma_\varepsilon = 4.00$<br>$\langle f_r \rangle = 11.30$ $\sigma_r = 11.30$<br>$\langle f_{inf} \rangle = 11.30$ $\sigma_{inf} = 11.30$<br>$\langle f_{tot} \rangle = 15.30$ $\sigma_{tot} = 11.99$ |
| $f_I$ Gamma Distribution: $\langle f_I \rangle = 5.80$ $\sigma_I = 3.02$ | | |

**Table 4:** Omicron variant distributions based on Refs. (30, 31, 38, 41). Meanings of quantities are as described in Table 2

| Omicron Variant | $f_\varepsilon$ Gamma Distribution | $f_\varepsilon$ Exponential Distribution |
| --- | --- | --- |
| $f_r$ Gamma Distribution | $\langle f_\varepsilon \rangle = 1.50$ $\sigma_\varepsilon = 1.00$<br>$\langle f_r \rangle = 12.50$ $\sigma_r = 1.77$<br>$\langle f_{inf} \rangle = 11.00$ $\sigma_{inf} = 2.03$<br>$\langle f_{tot} \rangle = 12.50$ $\sigma_{tot} = 1.77$ | $\langle f_\varepsilon \rangle = 1.50$ $\sigma_\varepsilon = 1.50$<br>$\langle f_r \rangle = 12.50$ $\sigma_r = 1.77$<br>$\langle f_{inf} \rangle = 11.00$ $\sigma_{inf} = 2.31$<br>$\langle f_{tot} \rangle = 12.50$ $\sigma_{tot} = 1.77$ |
| $f_r$ Exponential Distribution | $\langle f_\varepsilon \rangle = 1.50$ $\sigma_\varepsilon = 1.00$<br>$\langle f_r \rangle = 11.00$ $\sigma_r = 11.00$<br>$\langle f_{inf} \rangle = 11.00$ $\sigma_{inf} = 11.00$<br>$\langle f_{tot} \rangle = 12.50$ $\sigma_{tot} = 11.04$ | $\langle f_\varepsilon \rangle = 1.50$ $\sigma_\varepsilon = 1.50$<br>$\langle f_r \rangle = 11.00$ $\sigma_r = 11.00$<br>$\langle f_{inf} \rangle = 11.00$ $\sigma_{inf} = 11.00$<br>$\langle f_{tot} \rangle = 12.50$ $\sigma_{tot} = 11.10$ |
| $f_I$ Gamma Distribution: $\langle f_I \rangle = 3.00$ $\sigma_I = 1.22$ | | |

The remaining model parameters and values are summarized in Table 5. For all COVID variants, the PA and ES population fractions, *f*_*A*_ and *f*_*Y*_, respectively, are highly uncertain parameters, as estimates based on both modeling and clinical data place *f*_*A*_ anywhere from less than 1% to 90% (12, 13, 14, 15). Further, the value of *f*_*A*_ will depend precisely on our definition of the difference between the moderate to critical symptoms exhibited by the ES class versus the mild to no symptoms exhibited by the PA class. Evidence has suggested the majority of COVID cases to be mild (42, 43), so based on these observations, we choose *f*_*A*_ = 0.75 and *f*_*Y*_ = 0.25. Likewise, the relative values of the PA and ES transmission probabilities, *λ*_*A*_ and *λ*_*Y*_, respectively, are highly uncertain. Various studies have show that non-symptomatic individuals are as, or less, infectious than symptomatic individuals (13, 14), and that greater symptom severity correlates with higher viral loads (13, 14, 42). Further, a study on the close contacts of index cases has suggested symptomatic individuals to be more infectious than asymptomatic individuals (44). We therefore assume *λ*_*Y*_ = 2*λ*_*A*_. To determine absolute values, we assume an average contact rate of *β* = 4 per day and scale *λ*_*A*_ such that the model’s uncontrolled basic reproduction number under the exponential distribution assumptions (i.e. equivalent ODE model), denoted 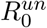, matches values taken from the literature (see the Supplementary Material section S1 and S2 for a derivation and discussion of the basic reproduction number). In other words, we define the following quantity:

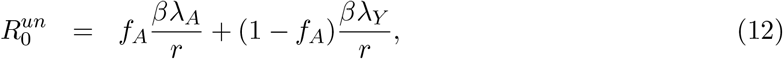

and then choose *λ*_*A*_ such that the above expression matches values for the different COVID variants, where the values of *f*_*A*_ and *β* are assumed, 1*/r* is equal to ⟨*f*_*inf*_ ⟩under the exponential distributions assumptions in Tables 2, 3, or 4, and where we assume *λ*_*Y*_ = 2*λ*_*A*_. Based on estimates of *R*_0_ during the initial phases of the pandemic (45, 46, 47), we set 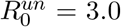 for the original COVID variant. Combining estimates from (11, 48, 49), we set 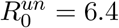 for the delta variant. Based on (49, 50), we set 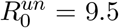 for the omicron variant. We note that changing exponential or gamma distribution assumptions for *f*_*r*_ and *f*_*ε*_ have a negligible impact on actual model basic reproduction number (see Supplementary Material section S2), so 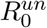 represents the uncontrolled basic reproduction number for all distribution and symptom onset assumptions.

**Table 5:** Model parameter definitions and numerical values used for COVID-19 variants.

| Parameter | Name | Meaning | Value | Refs |
| --- | --- | --- | --- | --- |
| $R_0^{un}$ | Uncontrolled basic reproduction number | Model $R_0$ with no testing or quarantine control | 3.0 (Original Variant)<br>6.4 (Delta Variant)<br>9.5 (Omicron Variant) | (11, 45, 46, 47, 48, 49, 50) |
| $\beta$ | Contact rate | Average number of contacts per individual per unit time | $4.0 \text{ (day)}^{-1}$ | Assumed |
| $\lambda_A$ | PA transmission probability | Probability of disease transmission per susceptible-permanently asymptomatic contact | 0.055 (Original Variant)<br>0.113 (Delta Variant)<br>0.173 (Omicron Variant) | Inferred from $R_0^{un}$ |
| $\lambda_Y$ | ES transmission probability | Probability of disease transmission per susceptible-eventually symptomatic contact | $2\lambda_A$ | (13, 14, 42, 44) |
| $f_A$ | Asymptomatic fraction | Fraction of infections which remain mild or asymptomatic | 0.75 | (12, 13, 14, 15, 42, 43) |
| $f_Y$ | Symptomatic fraction | Fraction of infections which become severe and symptomatic | $1 - f_A$ | - |
| $N$ | Population size | Total number of hosts (assumed fixed) | 50000 | Assumed |

#### 2.2.2 Generic diseases

In addition to the COVID-19 variants, we consider a generic disease similar to the original variant but with a variable offset between the mean incubation and latent period. Defining *z* = ⟨*f*_*I*_ ⟩ *−*⟨*f*_*ε*_ ⟩, we consider both positive and negative *z* values. Positive *z* values represent diseases for which symptom onset typically occurs after infectiousness onset, thus allowing for significant levels of presymptomatic transmission as occurs, for example, with the viruses SARS-CoV-2 and 2009 pandemic influenza H1N1 (51, 52, 28). Negative *z* values indicate viruses like SARS-CoV and MERS-CoV for which symptom onset typically occurs well before infectiousness onset or peak infectivity, thus making pre-symptomatic transmission insignificant or absent entirely (28, 29). In all cases, we assume fixed gamma distributions for *f*_*ε*_ and *f*_*r*_ similar to those of the original COVID-19 variant and vary *z* by considering different gamma distributed incubation periods *f*_*I*_. Assumed means and standard deviations for *f*_*ε*_, *f*_*r*_, and *f*_*I*_ for the specific *z* values under consideration are given in Table 6, and the distributions are pictured graphically in Fig. 4. Remaining model parameters are taken to be equivalent to those of COVID-19 in Table 5, where we will consider a variety of 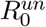 and corresponding *λ*_*A*_ values.

**Table 6:** Gamma distribution parameters for generic diseases with different offsets between mean incubation and latent periods denoted by *z* = ⟨*f*_*I*_ ⟩ *−* ⟨*f*_*ε*_ ⟩. Means ⟨ ⟩, standard deviations *σ*, and *z* values are given in units of days.

| Offset | $z = -4.5$ | $z = -3.0$ | $z = -1.5$ | $z = 0$ | $z = 1.5$ | $z = 3.0$ | $z = 4.5$ |
| --- | --- | --- | --- | --- | --- | --- | --- |
| $\langle f_I \rangle$ | 1.50 | 3.00 | 4.50 | 6.00 | 7.50 | 9.00 | 10.50 |
| $\sigma_I$ | 1.22 | 1.91 | 2.27 | 2.83 | 2.83 | 2.83 | 2.83 |
| $\langle f_\varepsilon \rangle = 6.00, \sigma_\varepsilon = 2.83 \quad \langle f_r \rangle = 17.00, \sigma_r = 2.06$<br>$\langle f_{inf} \rangle = 11.02, \sigma_{inf} = 3.45 \quad \langle f_{tot} \rangle = 17.02, \sigma_{tot} = 2.06$ | | | | | | | |

**Figure 4.**
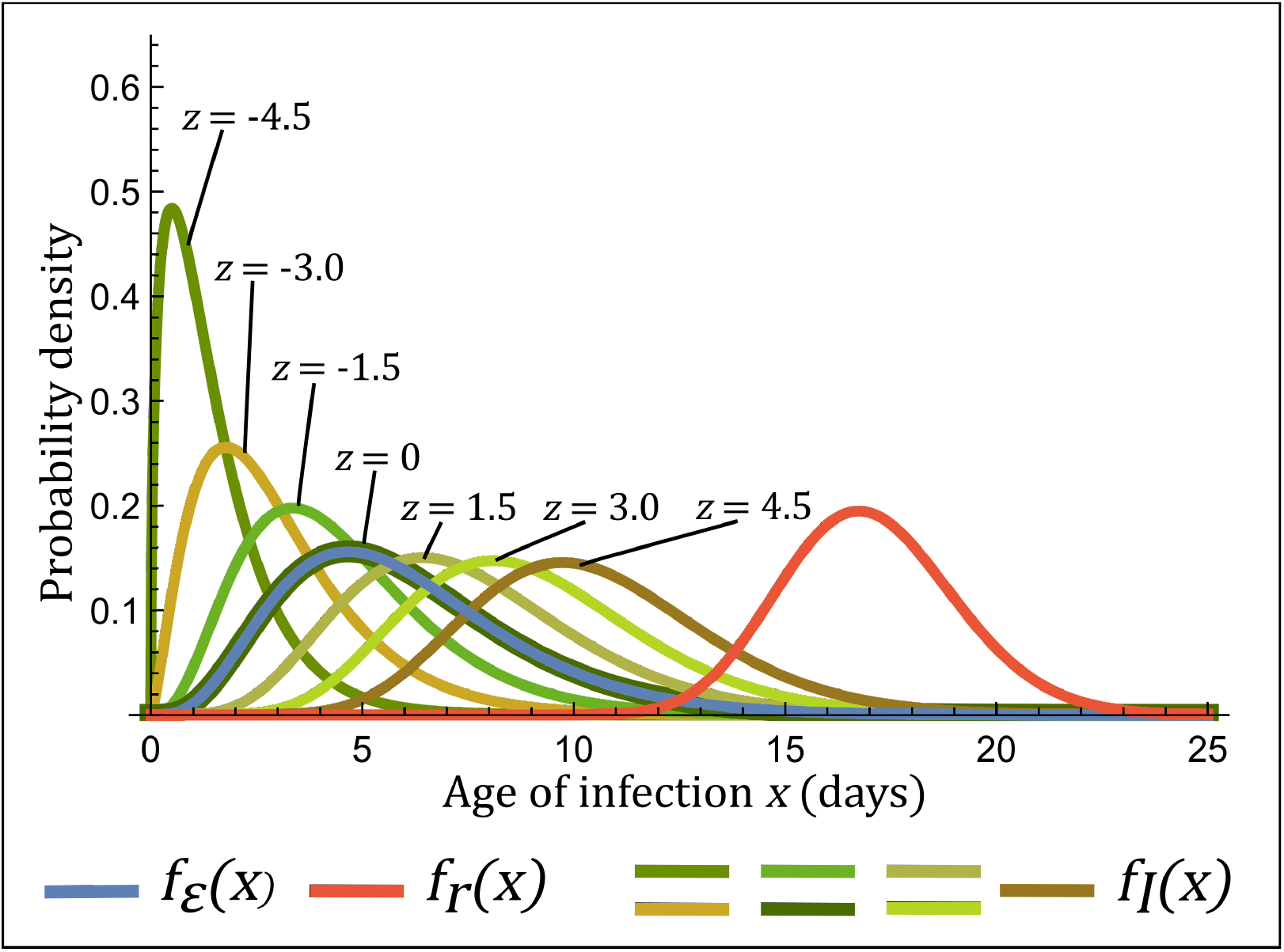
Latent period distribution *f*_*ε*_(*x*), incubation period distributions *f*_*I*_(*x*), and post-infectiousness onset recovery day distribution *f*_*r*_(*x*) as functions of the age of infection *x* for generic diseases with different offsets *z* between mean incubation and latent periods indicated in units of days.

### 2.3 Numerical integration and optimization

We compute the discretized dynamical equations for the system (7) using the upwind scheme for non-linear partial integro-differential equations with integral boundary conditions (53), and then use the midpoint method to integrate the system forward in time. We assume initial infection conditions *e*_*A*_(0, *x*)Δ*x* = *f*_*A*_ and *e*_*Y*_ (0, *x*)Δ*x* = *f*_*Y*_ for *x* ∈ [0, Δ*x*], where Δ*x* = 1*/*6 days is the discretization length for the age of infection domain, and *e*_*A*_(0, *x*) = *e*_*A*_(0, *y*) = 0 for *x >* Δ*x*. The system is assumed otherwise to be initially completely susceptible such that *S*(0) = *N* − 1 and *a*(0, *x*) = *y*(0, *x*) = *q*(0, *x*) = *R*(0) = *U* (0) = 0 for all *x*. Age of infection integrals are computed using the trapezoid rule where we assume a finite domain with an upper bound *x*_*max*_ = 80 days (the number of infected individuals who would otherwise recover after *x*_*max*_ is negligible for all disease variants considered). We integrate the model equations from an initial time *t*_0_ = 0 to final time *t*_*f*_ = 2 years using a time step Δ*t* = 1/10 days. When implementing testing and quarantine control, we find optimal allocation strategies *ρ* for reducing the total infection size *S*(*t*_0_) − *S*(*t*_*f*_) under a variety of *C, η*, and *κ* values using the *fmincon* function in *Matlab R2021b*.

## 3 Results

### 3.1 Optimal total infection size reduction and disease controllability

Figures 6 and 7 display total infection sizes under optimal resource allocation strategies as a function of testing capacity for the different COVID variants as well as the generic disease with *z* and 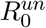 comparable to that of delta and omicron variants (*z* = 1.5 days and 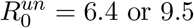). Curve color and dashing pattern represent different combinations of assumptions regarding symptom onset (incubation symptoms or correlated symptoms) and distributions for *f*_*ε*_ and *f*_*r*_ (gamma or exponential) as detailed by the legend in Fig. 5. Different plots represent different disease variants and different choices for *η* and *κ* values. The presented results focus on two particular cases: “random testing” (*κ* = 1.00, *η* = 0) and “realistic testing” (*κ* = 0.85, *η* = 0.75). Random testing represents a population-wide pure random sampling non-clinical testing program, while realistic testing represents a non-clinical testing scenario more likely to be encountered in the real world, where contact tracing and natural biases result in increased testing of infected individuals but where some of the population is inaccessible or resistant to testing efforts. The case of no control is represented at zero testing capacity (*C* = 0). By definition, symptom onset assumptions and testing parameter values have no influence on model outcomes at *C* = 0, but interestingly, we see that exponential versus gamma distribution assumptions for *f*_*ε*_ and *f*_*r*_ have no meaningful impact on total infection size at *C* = 0 even though model dynamics differ.

**Figure 5.**
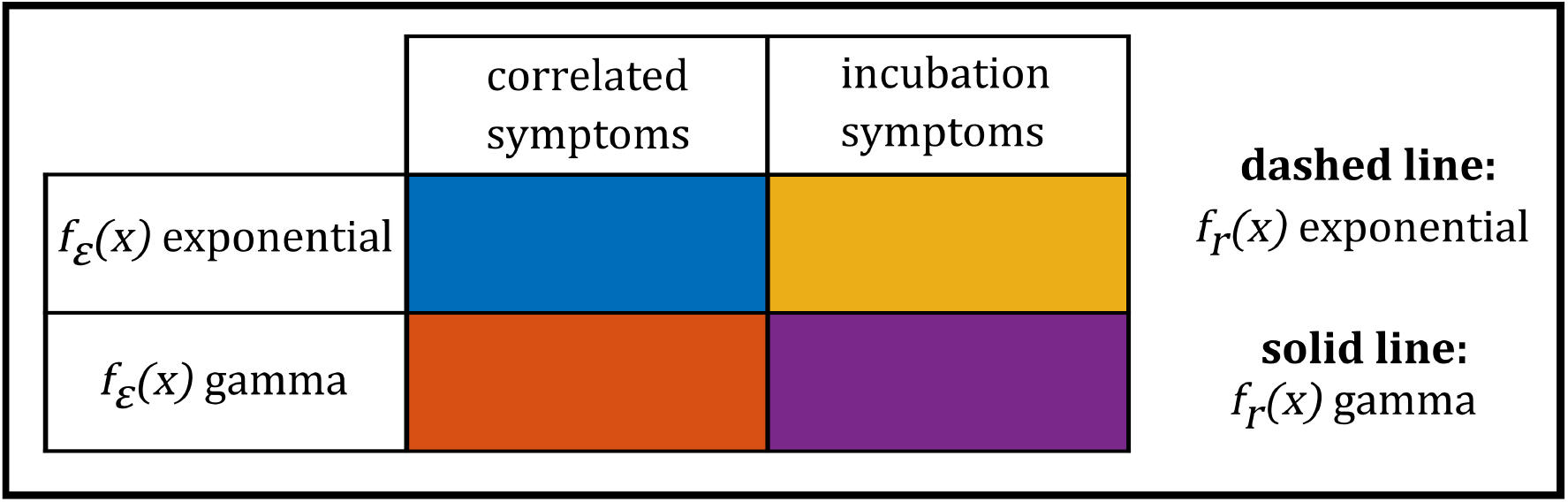
Legend for interpreting curve color and style in Figs. 6, 7, 8, 9, and 10. Different colors represent different combinations of exponential and gamma distribution assumptions for *f*_*ε*_ along with different assumptions for correlated versus incubation symptom onset as indicated by the colored table. Dashed lines indicate the exponential distribution assumption for *f*_*r*_ while solid lines indicate the gamma distribution assumption. The model with all IPDE elements is given by a solid purple curve while the ODE equivalent model is given by a dashed blue curve.

**Figure 6.**
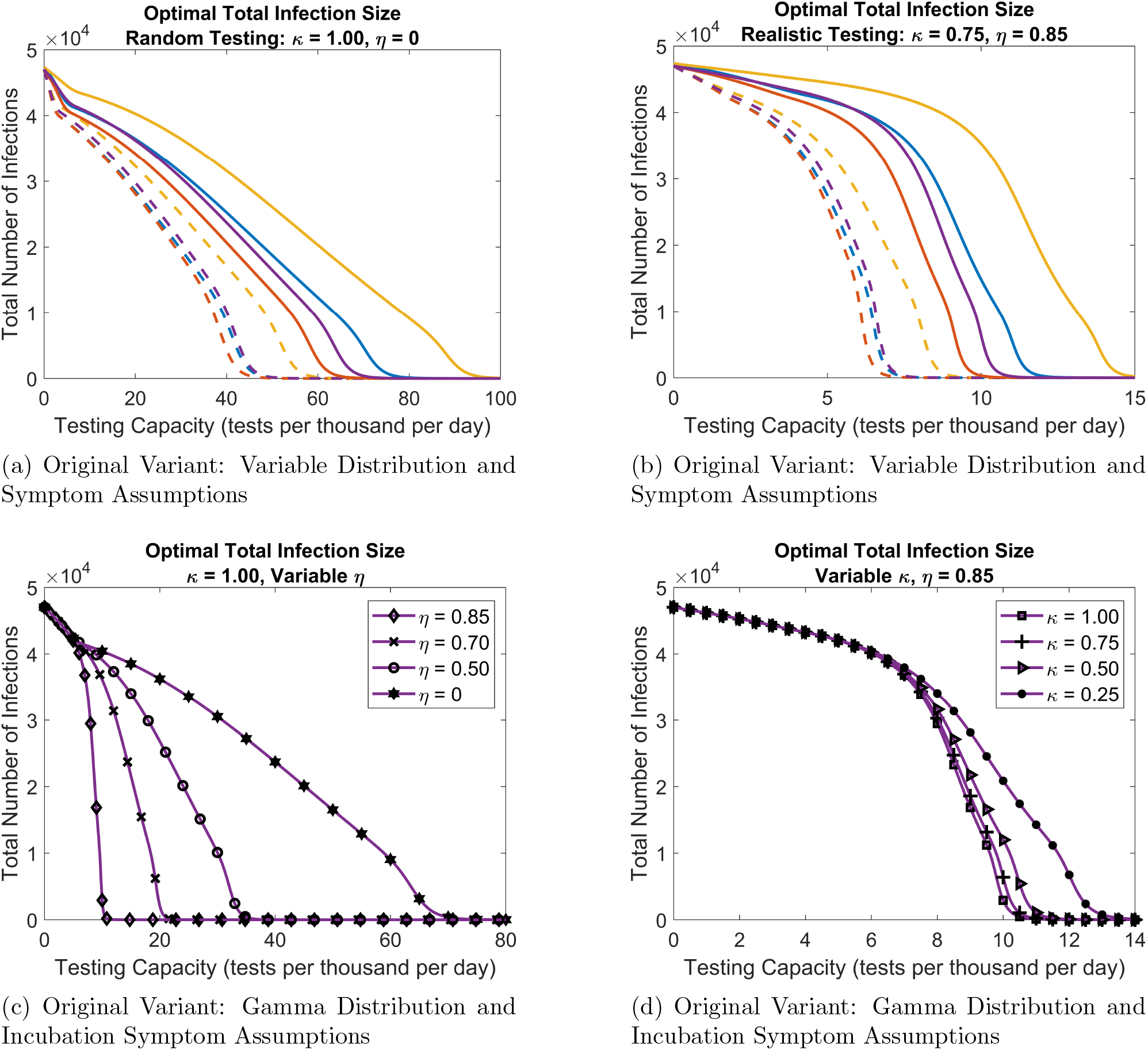
Total infection sizes under optimal allocation strategies as a function of testing capacity for the original COVID variant under a variety of symptom onset, period distribution, and testing parameter assumptions. The meaning of curve color and dashed versus solid curves is given in Fig. 5. Note the changes in x-axis scale for each plot.

**Figure 7.**
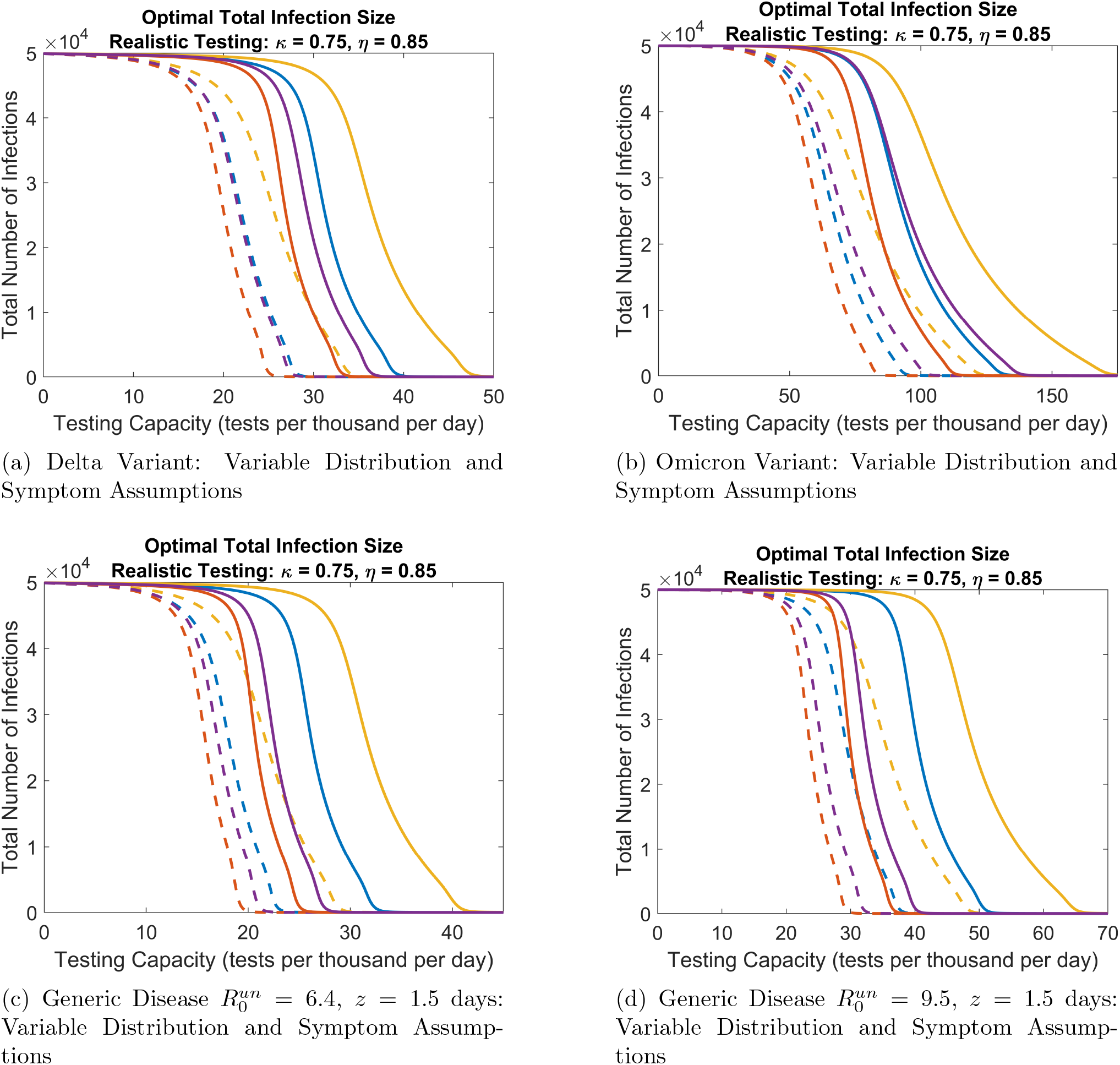
Total infection sizes under optimal allocation strategies as a function of testing capacity for the delta and omicron COVID variants, as well a generic disease similar to COVID-19, under a variety of symptom onset, period distribution, and testing parameter assumptions. The meaning of curve color and dashed versus solid curves is given in Fig. 5. The generic disease assumes 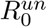 values equal to those of the delta and omicron COVID variants with a mean latent period ⟨*f*_*ε*_ ⟩ = 6.00 days comparable to that of the original variant, and assumes a incubation-latent offset *z* = 1.5 days similar to all COVID variants. Note the changes in x-axis scale for each plot.

Generally, as the testing capacity *C* increases, Figs. 6 and 7 show that the total infection size decreases when resources are distributed optimally. In particular, complete disease eradication is possible at reasonably achievable testing capacities for a less infectious variant when non-clinical testing can be effectively targeted at individuals who are actually infected (large *η* in Figs. 6b and 6d). However, if non-clinical testing does not target infected individuals (small *η* in Figs. 6a and 6c), or if the variant is of greater infectiousness (Fig. 7), eradication may not be achievable at the reduced testing capacities available during the initial phases of a novel disease outbreak. The accessibility *κ* has a smaller influence on reducing total infections compared to the concentration *η*, and disease eradication may possible even for smaller accessibility levels (Fig. 6d). This result emphasizes the importance of implementing effective contact tracing programs for COVID-like diseases even if such programs can only reach a relatively small number of individuals.

Together, Figs. 6 and 7 exemplify the influences of symptom onset assumptions, period distribution assumptions, testing parameter values, and disease variant characteristics on disease controllability. By controllability, we specifically mean the amount of testing capacity required to achieve a given reduction in total infection size. We consider a set ‘A’ of assumptions and parameters to be less controllable than another set ‘B’ if set ‘A’ requires a larger testing capacity to reduce total infection size to half, for example, of the uncontrolled value under optimal resource allocation strategies (see Supplementary Material section S4 for alternative but equivalent measures of controllability involving the basic reproduction number).

Gamma versus exponential distribution assumptions for *f*_*r*_ have a profound effect on controllability. Comparing a dashed curve (exponential) to a solid curve (gamma) of the same color in either Fig. 6a, 6b, or 7 shows that the gamma assumption produces a large decrease in controllability relative to the exponential assumption. On the other hand, the gamma assumption for *f*_*ε*_ increases controllability relative to the exponential assumption. This is seen in Fig. 6a, 6b, or 7 by comparing a solid orange curve to a solid blue curve or a solid purple curve to a solid gold curve, and likewise for dashed curves. The effects of incubation symptom versus correlated symptom assumptions are exemplified by comparing a solid gold to solid blue curve or solid purple to solid orange curve, and likewise for dashed curves. Here, incubation symptom assumptions decrease controllability relative to correlated symptom assumptions. Interestingly, for a given assumption on *f*_*r*_, the decrease in controllability due to incubation symptoms is nullified to some extent by the increase in controllability due to a gamma distributed *f*_*ε*_ such that solid (dashed) blue and purple curves can lie close to one another bounded by solid (dashed) red and gold curves. In other words, the IPDE specific assumptions for latent period length and symptom onset tend to counteract one another. However, the full IPDE model (solid purple curve) generally shows a significant reduction in controllability relative to the full ODE model (dashed blue curve).

The influences of the testing parameters *η* and *κ* on controllability are shown in Figs. 6c and 6d, where smaller values tend to decrease controllability. The influences of disease variant characteristics are seen by comparing Fig. 6b to the corresponding curves in all plots in Fig. 7. Here, the omicron COVID variant is significantly less controllable than the delta variant, and the delta variant is moderately less controllable than the original variant. These controllability reductions may be caused by either increases in transmissibility or decreases in the mean latent period length associated with each COVID variant, where 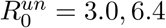, and 9.5 and ⟨*f*_*ε*_ ⟩ = 5.48, 4.00, and 1.50 days for the original, delta, and omicron variants, respectively (the mean incubation-latent offsets *z* and mean infectious period lengths are approximately equal for all variants). The independent influences of transmissibility and latent period length can be deduced using the generic disease in Figs. 7c and 7d. Here, the mean latent period ⟨*f*_*ε*_⟩ = 6.00 days is comparable to that of the original variant, and comparing Fig. 6b to Figs. 7c and 7d thus shows that increases in transmissibility alone cause decreases in controllability. Likewise, comparing Fig. 7a and 7b to Fig. 7c and Fig. 7d, respectively, shows that decreases in ⟨*f*_*ε*_ ⟩ alone cause decreases in controllability. A summary of all observed controllability reducing factors is given in Table 7.

**Table 7:** List of controllability reducing factors observed in Figs. 6 and 7 for COVID variants and generically parameterized diseases with incubation-latent offset *z* = 1.5 days.

| Controllability reducing factors for COVID-like diseases |
| --- |
| $f_r$ gamma distribution |
| $f_\varepsilon$ exponential distribution |
| reduction in non-clinical concentration $\eta$ |
| reduction in non-clinical accessibility $\kappa$ |
| increase in overall transmissibility (i.e. $R_0^{un}$) |
| reduction in mean latent period $\langle f_\varepsilon \rangle$ |
| incubation symptoms |

### 3.2 Optimal allocation strategies

Figures 8 and 9 show the optimal resource allocation strategies *ρ* as a function of testing capacity *C* corresponding to the total infection size reductions in Figs. 6 and 7. Curve colors and styles are interpreted analogously according to the legend in Fig. 5. The value *ρ* = 0 represents a clinicaltesting only strategy while *ρ* = 1 represents a non-clinical testing only strategy, with intermediate values represent mixed clinical and non-clinical testing strategies. In all cases, we find that optimal strategies call for clinical testing only at low testing capacities up to some strategy threshold capacity *C*^*th*^, beyond which optimal strategies become mixed. As testing capacity increases further beyond *C*^*th*^, optimal strategies call for greater and greater shares of resources to be devoted to non-clinical testing.

**Figure 8.**
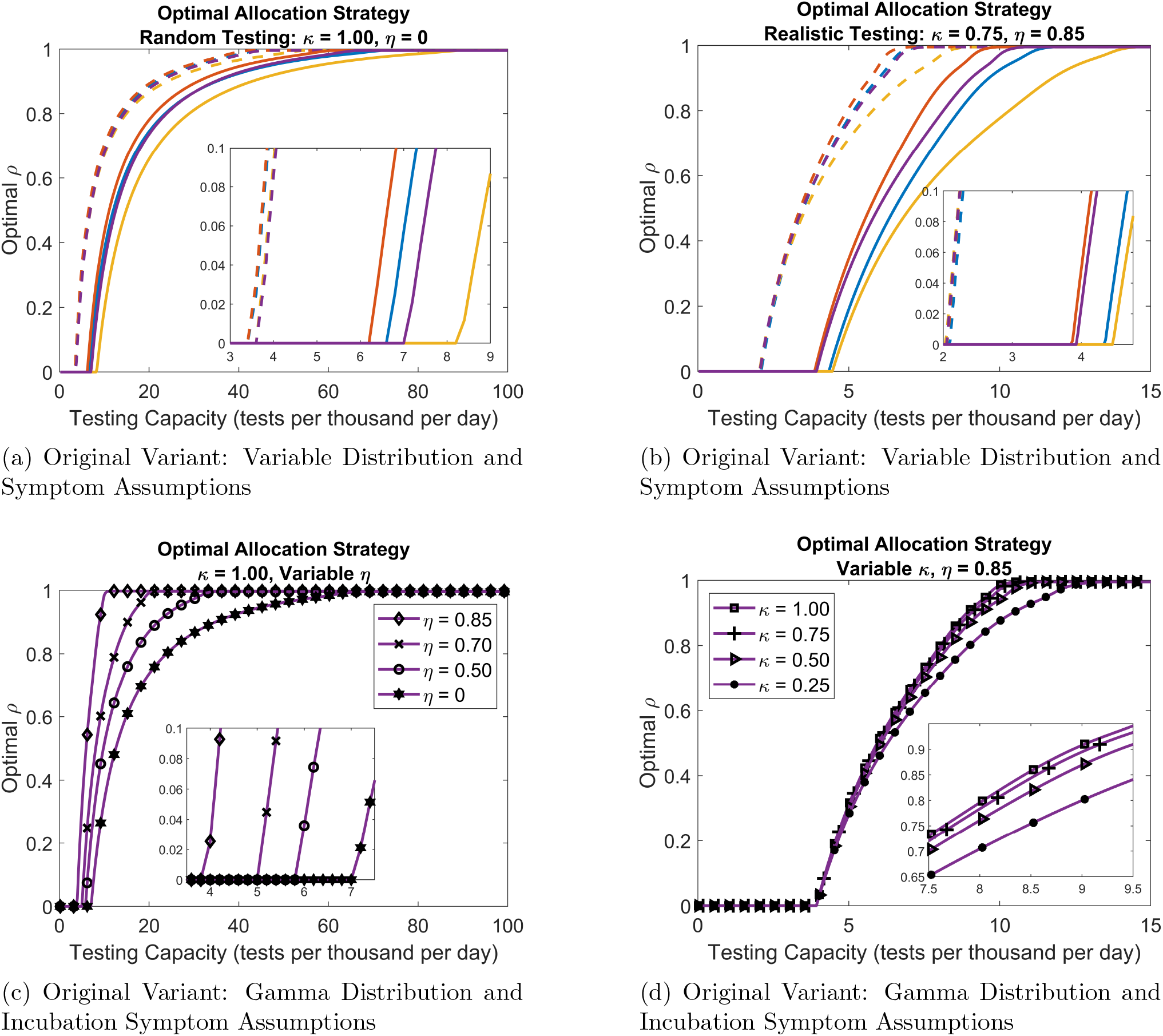
Optimal resource allocation strategies for the original COVID variant corresponding to the total infection sizes in Fig. 6. The value *ρ* = 0 corresponds to clinical-testing only strategy, *ρ* = 1 corresponds to non-clinical testing only, with intermediate values representing mixed strategies. Insets within plots (aside from Fig. 8d) highlight strategy threshold testing capacity values *C*^*th*^ where strategies switch from clinical only to mixed. Note the changes in x-axis scale for each plot.

**Figure 9.**
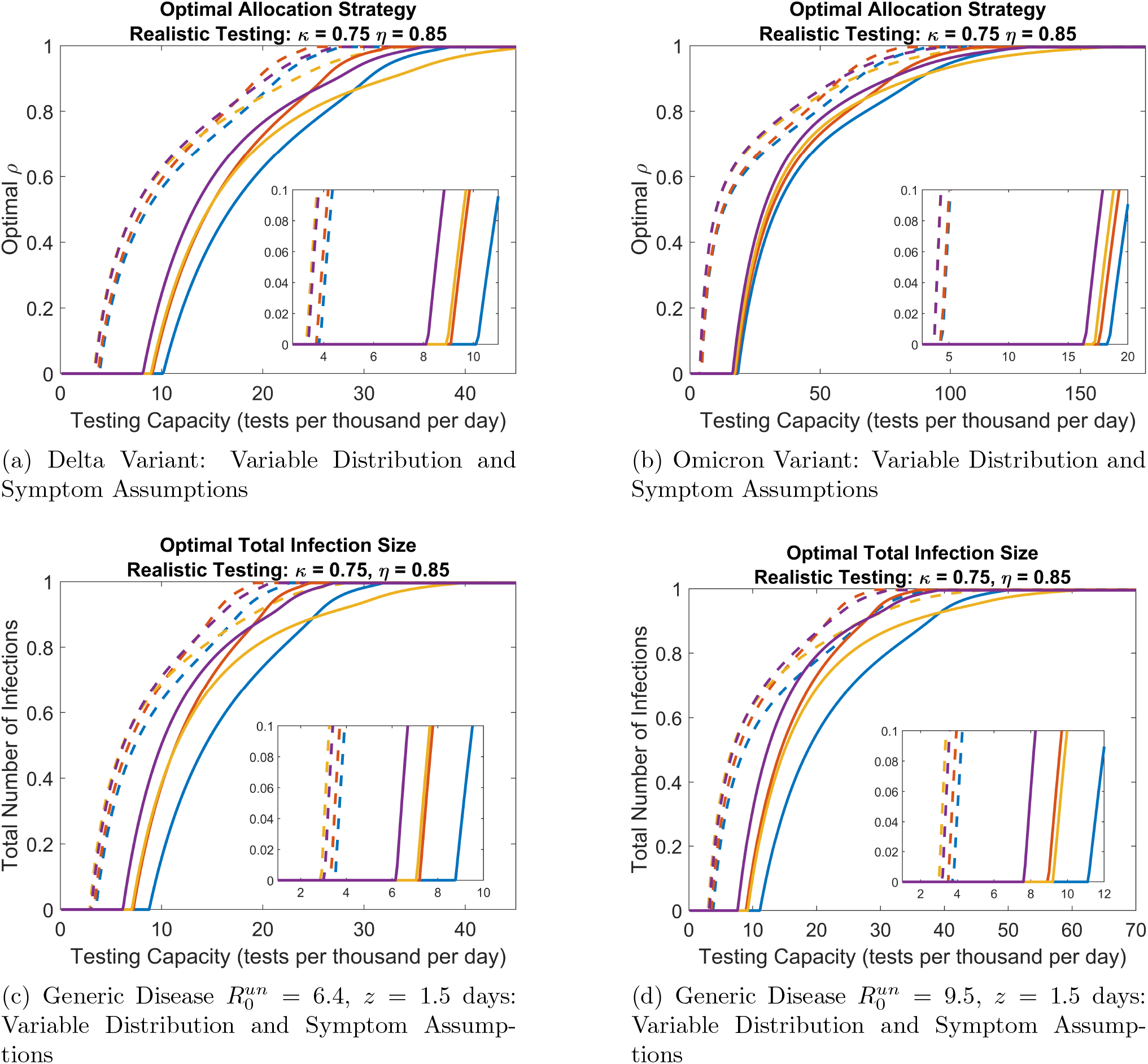
Optimal resource allocation strategies for the delta and omicron COVID variants as well as comparable generic diseases with a longer mean incubation period ⟨*f*_*ε*_ ⟩ = 6.0 days. Optimal strategies correspond to the optimal total infection sizes in Fig. 7. The value *ρ* = 0 corresponds to clinical-testing only strategy, *ρ* = 1 corresponds to non-clinical testing only, with intermediate values representing mixed strategies. Insets within plots highlight strategy threshold testing capacity values *C*^*th*^ where strategies switch from clinical only to mixed. Note the changes in x-axis scale for each plot.

The insets in each plot (except Fig. 8d) zoom in on *C*^*th*^ values for the various distribution, symptom, testing parameter, and disease parameter assumptions. Figures 8 and 9 show that all of these factors except the non-clinical accessibility *κ* are important in determining *C*^*th*^, while Fig. 8d indicates that *C*^*th*^ is in fact independent of *κ*. This observation can be explained from our model equations, where the total rate of flow of infected individuals to the quarantine class due to non-clinical testing is given by 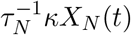. At the threshold *C*^*th*^, the optimal fraction of resources *ρ* devoted to nonclinical testing is only infinitesimally larger than zero, and Eq. (5) implies 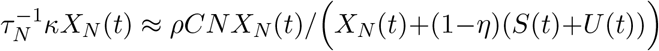 for very small *ρ*. We thus conclude that the rate at which individuals are quarantined by optimally allocated non-clinical testing is independent of *κ* at testing capacities near *C*^*th*^, and that *κ* effectively vanishes from the model equations under optimal strategies at *C*^*th*^.

### 3.3 Controllability and the role of non-clinical testing

Figures 6, 7, 8, and 9 together depict the relationship between controllability and the role of non-clinical testing in optimal strategies. We say that the role of non-clinical testing is increased for set ‘A’ of distribution, symptom onset, testing parameter, and disease parameter assumptions relative to another set ‘B’ if the optimal *ρ* value at a given testing capacity for set ‘A’ is larger. Applying the same comparative analyses to Figs. 8 and 9 as was done for Figs. 6 and 7 in Sec. 3.1, we find that, except for the incubation symptoms assumption, all factors that reduce controllability in Table 7 coincide with the factors that reduce the role of non-clinical testing. Thus there exists a strong relationship between controllability influencing factors and the role of non-clinical testing in optimal strategies.

The relationship between non-clinical testing and symptom onset assumptions is more nuanced than for the other controllability reducing factors. Understanding this relationship is important for disease control, as symptom onset assumptions either incorporate (incubation symptoms) or disregard (correlated symptoms) the capacity for presymptomatic transmission associated with the incubation-latent period offset for COVID-19. In some cases (compare gold to blue or purple to orange curves in Figs. 8a and 8b), incubation symptoms decrease the role of non-clinical testing relative to correlated symptoms, thus following the pattern of controllability reducing factors coinciding with non-clinical testing reducing factors. In other cases (Fig. 9), incubation symptoms increases the role of non-clinical testing for a large range of testing capacities above the strategy thresholds *C*^*th*^ even though controllability is reduced. Of further complication, in these cases, there exist smaller ranges of larger testing capacities where the role of non-clinical testing is increased by the incubation symptoms assumption.

We utilize strategy testing capacity thresholds *C*^*th*^ to analyze the relationship between non-clinical testing and symptom onset assumptions. Specifically, in Figs. 8a and 8b, the reduced roles of non-clinical testing under the incubation symptom assumptions correspond to larger *C*^*th*^ values in comparison to the correlated symptoms assumptions. Likewise, in Fig. 9, the enhanced roles of non-clinical testing over large ranges of testing capacities correspond to smaller *C*^*th*^ for incubation symptoms in comparison to correlated symptoms. Thus, increases and decreases in *C*^*th*^ due to symptom onset assumptions are simple indicators of decreases and increases, respectively, in the roles of non-clinical testing. To further simplify our analysis, we restrict our attention to the most realistic model assumptions using gamma distributions for *f*_*ε*_ and *f*_*r*_ to compare incubation symptoms (solid purple curves) to correlated symptoms (solid orange curves).

Numerically calculating *C*^*th*^ values for a spectrum of *η* values, we plot the results as curves in the (*C, η*) plane, where different curves represent different variant and symptom onset assumptions (Fig. 10). For a given variant, if the correlated symptom curve (orange) falls to the left of the incubation symptom (purple) curve at a given *η*, then *C*^*th*^ is larger for incubation symptoms, thus implying that incubation symptoms reduce the role of non-clinical testing. Conversely, if the correlated symptom curve falls to the right of the incubation symptoms curve, incubation symptoms enhance the role of non-clinical testing. Further, for each variant, there exists a threshold concentration *η*^*th*^ (indicated by black circles in Fig. 10), above which the role of non-clinical testing is always enhanced, and below which it is always reduced, where threshold values decrease as variant strength increases. Thus, whether or not the presence of presymptomatic transmission warrants allocation of additional resources to non-clinical testing than would otherwise be optimal depends on both variant strength and the precision to which non-clinical tests can locate infected individuals: variants with greater infectiousness require less precision to justify additional resources for non-clinical testing.

**Figure 10.**
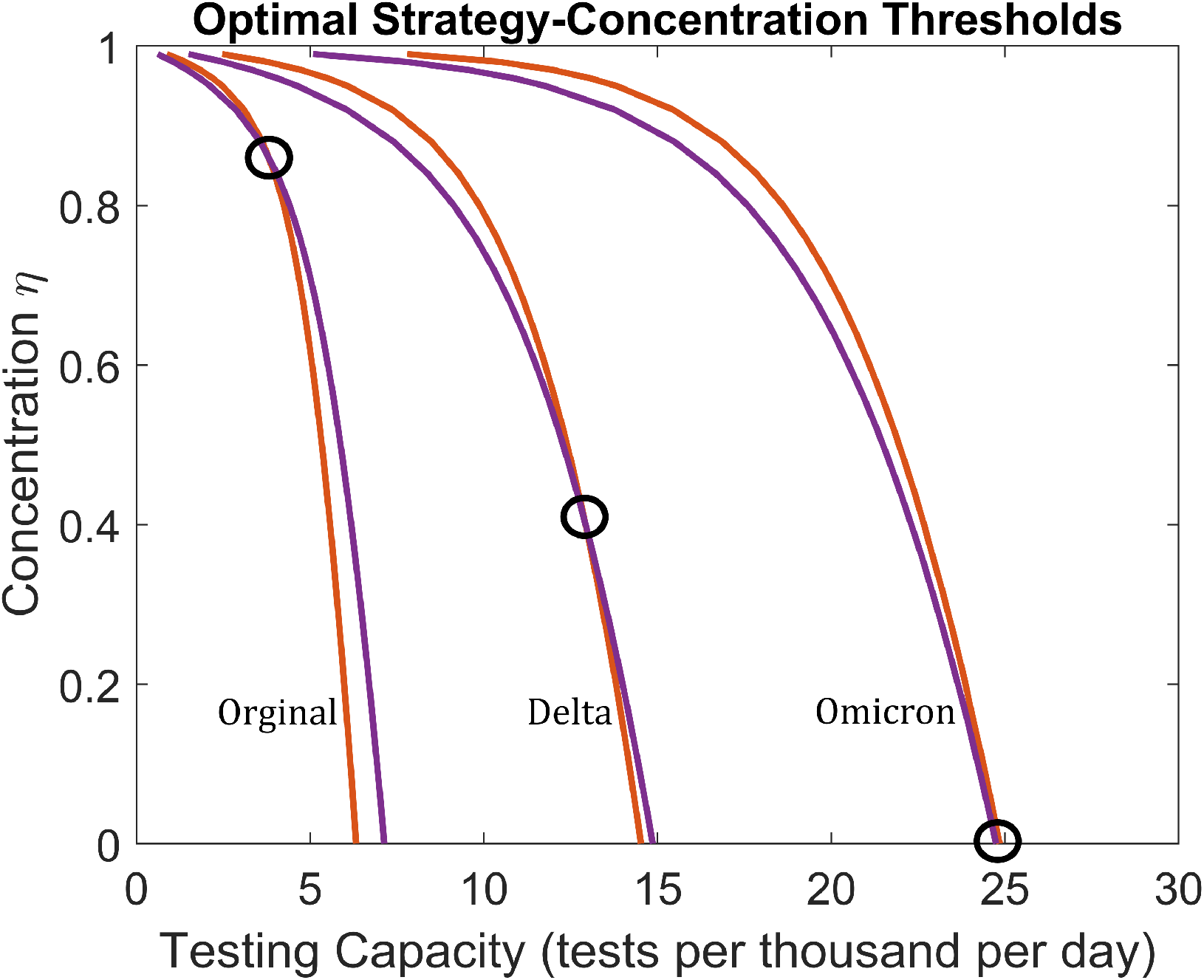
Strategy threshold testing capacities at various concentration levels *η* for the different COVID variants. Curves represent (*C, η*) values where the strategy threshold capacity *C*^*th*^ occurs, assuming either incubation symptoms (purple) or correlated symptoms (orange) with gamma distributed *f*_*r*_ and *f*_*ε*_. At (*C, η*) points above or to the right of a given curve, optimal strategies are mixed clinical and non-clinical for that variant and symptom assumption. At points below or to the left, optimal strategies are clinical only. Circles indicate points where *C*^*th*^ is equivalent for both symptom assumptions. At *η* values above a circle, incubation symptoms increases the role of non-clinical testing in optimal strategies for the corresponding variant. At *η* values below a circle, incubation symptoms reduce the role of non-clinical testing.

Figure 10 is of additional value as a practical result for disease management officials when determining whether or not complicated resource allocation decisions need be considered. To see this, suppose that the available testing capacity *C* and an estimate for the concentration *η* of a non-clinical testing program are known. Then, if the corresponding (*C, η*) value falls to the left of or below a variant curve in Fig. 10, the optimal strategy for that variant is clinical testing only, and officials need not consider difficult choices in allocating resources. Otherwise, if the (*C, η*) value falls to the right of or above a variant curve, the optimal strategy is mixed clinical and non-clinical testing, and officials know that critical decisions must be made to ensure that resources are properly allocated.

### 3.4 Variable incubation-latent offsets

In Fig. 11, we plot optimal total infection sizes and corresponding optimal resource allocation strategies for the generic disease with 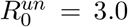 under a variety of incubation-latent offsets *z*. Here, we consider only gamma distributions for *f*_*r*_ and *f*_*ε*_, and we consider both incubation and correlated symptom assumptions (the offset *z* is irrelevant under correlated symptoms). The offset *z* is found to have a strong influence on controllability (Figs. 11a and 11b); as *z* increases from negative to positive values, controllability decreases significantly. Thus, diseases for which symptom onset typically occurs before peak infectiousness are more easily controlled with symptom-based interventions compared to diseases for which symptom onset typically occurs after infectiousness onset (i.e. presymptomatic transmission). Further, relative to the correlated symptoms assumption, incubation symptoms are more controllable for *z* < 0 and less controllable for *z* > 0 (Figs. 11a and 11b). This finding agrees with the notion that diseases for which symptoms precede significant infectiousness are easier to control; if a model disregards the offset between symptom and infectiousness onset, it will underestimate controllability for diseases with *z* < 0 and overestimate controllability for diseases with *z >* 0. Thus, Table 1 should be modified to state that incubation symptoms are a controllability reducing factor only for diseases with *z* > 0, while correlated symptoms are a controllability reducing factor for *z* < 0. This expanded list of controllability reducing factors is given in Table 8. For the case *z* = 0, controllability under incubation symptoms is close to (but not exactly equal to) controllability under correlated symptoms (Figs. 11a and 11b). Apparently, when symptom and infectiousness onset occur together only on average rather than always together, controllability is very slightly reduced.

**Figure 11.**
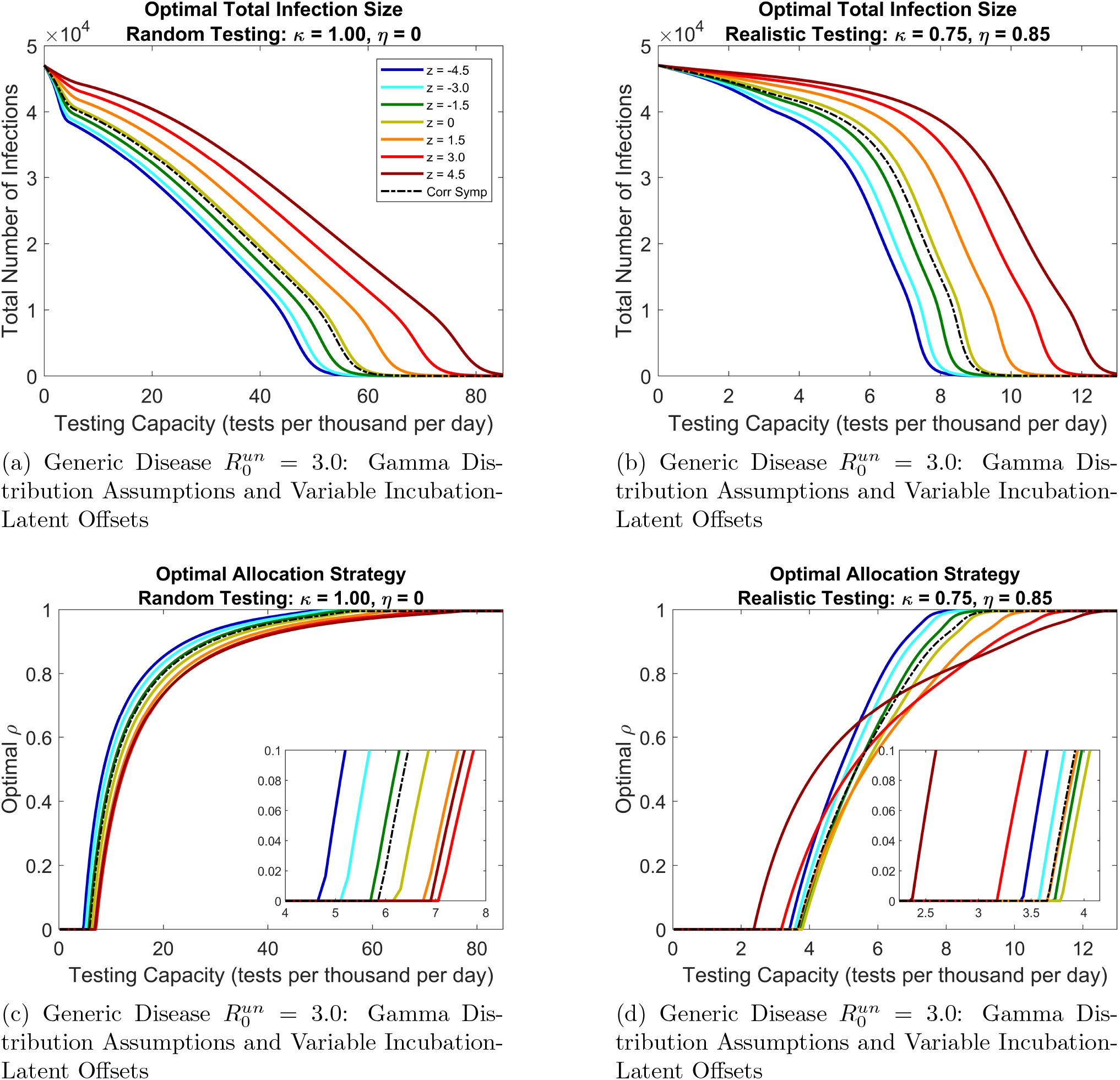
Optimal total infection sizes and corresponding optimal resource allocation strategies for the generic disease with 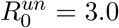, assuming gamma distributions for *f*_*ε*_ and *f*_*r*_. Curve colors represent different offsets *z* = ⟨*f*_*I*_ ⟩ − ⟨*f*_*ε*_ ⟩ between mean incubation and latent periods as indicated by the legend in Fig. 11a (measured in units of days). Black dashed curves represent the correlated symptoms assumption where *z* values are irrelevant. Insets in Figs. 11c and 11d zoom in on strategy threshold testing capacities *C*^*th*^ where optimal strategies switch from clinical only to mixed clinical and non-clinical.

**Table 8:** Expanded list of controllability reducing factors for both *z* > 0 and *z* < 0 diseases as observed in Figs. 6, 7, and 11.

| **Controllability reducing factors** |
| --- |
| $f_r$ gamma distribution |
| $f_\varepsilon$ exponential distribution |
| reduction in non-clinical concentration $\eta$ |
| reduction in non-clinical accessibility $\kappa$ |
| increase in overall transmissibility (i.e. $R_0^{un}$) |
| reduction in mean latent period $\langle f_\varepsilon \rangle$ |
| incubation symptoms when $z > 0$ |
| correlated symptoms when $z < 0$ |

As a function of testing capacity, optimal strategies for all *z* behave qualitatively similar to those of the COVID-19 variants; optimal strategies are clinical-only at low testing capacities and become mixed clinical and non-clinical beyond a threshold capacity *C*^*th*^ (Figs. 11c and 11d). As with the COVID-19 variants, the relationships between symptom onset assumptions, controllability, and optimal allocation strategies are complicated for the generic disease. For random testing (Fig. 11c), the role of non-clinical testing decreases (as measured by increases in *C*^*th*^) as *z* increases in the interval [*−*4.5, 1.5], thus following the trend of controllability reducing factors coinciding with non-clinical testing reducing factors. However, the role of non-clinical testing increases for *z* = 4.5 days relative to *z* = 1.5 days even though controllability decreases. For realistic testing (Fig. 11d), the role of non-clinical testing decreases progressively as *z* increases in the interval [*−*4.5, 0], after which it progressively increases as *z* increases in [0, 4.5]. Of further complication, for larger 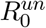 values (see Supplementary Material Figs. S10 and S11), the role of non-clinical testing can increase progressively as *z* increases despite the progressive decrease in controllability.

The role of non-clinical testing for incubation symptoms relative to that of correlated symptoms displays distinct behavior for *z* < 0 and *z* ≥ 0. Specifically, for *z* = −1.5 days (green curve in Figs. 11c and 11d), the role of non-clinical testing is enhanced relative to correlated symptoms at *η* = 0, but is reduced at *η* = 0.85. This behavior is counterintuitive; one would expect that if non-clinical testing is effective enough at low *η* to have an enhanced role in optimal strategies, then it would also have an enhanced role at larger *η*, as increasing *η* would increase its efficacy further. This is indeed the case for the COVID-19 variants that have a positive *z* approximately equal to 1.50 days (Fig. 10). For these variants, the role of non-clinical testing becomes enhanced relative to correlated symptoms at some threshold concentration *η*^*th*^, and then remains enhanced for all *η* > *η*^*th*^. Figure 12 presents the analog to Fig. 10 for the generic disease for 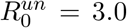 (see Supplementary Material Figs. 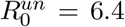 and 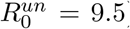. Here, we numerically calculate the strategy threshold capacities *C*^*th*^ for a spectrum of *η* values for each *z* offset and the correlated symptoms assumptions, and then plot the results as curves in the (*C, η*) plane. For each *z* curve, there is a threshold concentration *η*^*th*^ where a crossing with the correlated symptoms curve occurs. *z ≥* 0 curves are to the right of the correlated symptoms curve for *η < η*^*th*^ (indicating diminished non-clinical testing roles) and to the left for *η* > *η*^*th*^ (indicating enhanced non-clinical testing roles). However, *z <* 0 curves are to the left of the correlated symptoms curve for *η* < *η*^*th*^ (indicating enhanced non-clinical testing roles) and to the right for *η* > *η*^*th*^ (indicating diminished non-clinical testing roles). We thus find major qualitative differences between *z* > 0 diseases (i.e. those with presymptomatic transmission) and *z* < 0 diseases (i.e those with symptom onset before infectiousness) in regards to the influence of realistic incubation and latent periods on optimal resource allocation strategies relative to simpler models.

**Figure 12.**
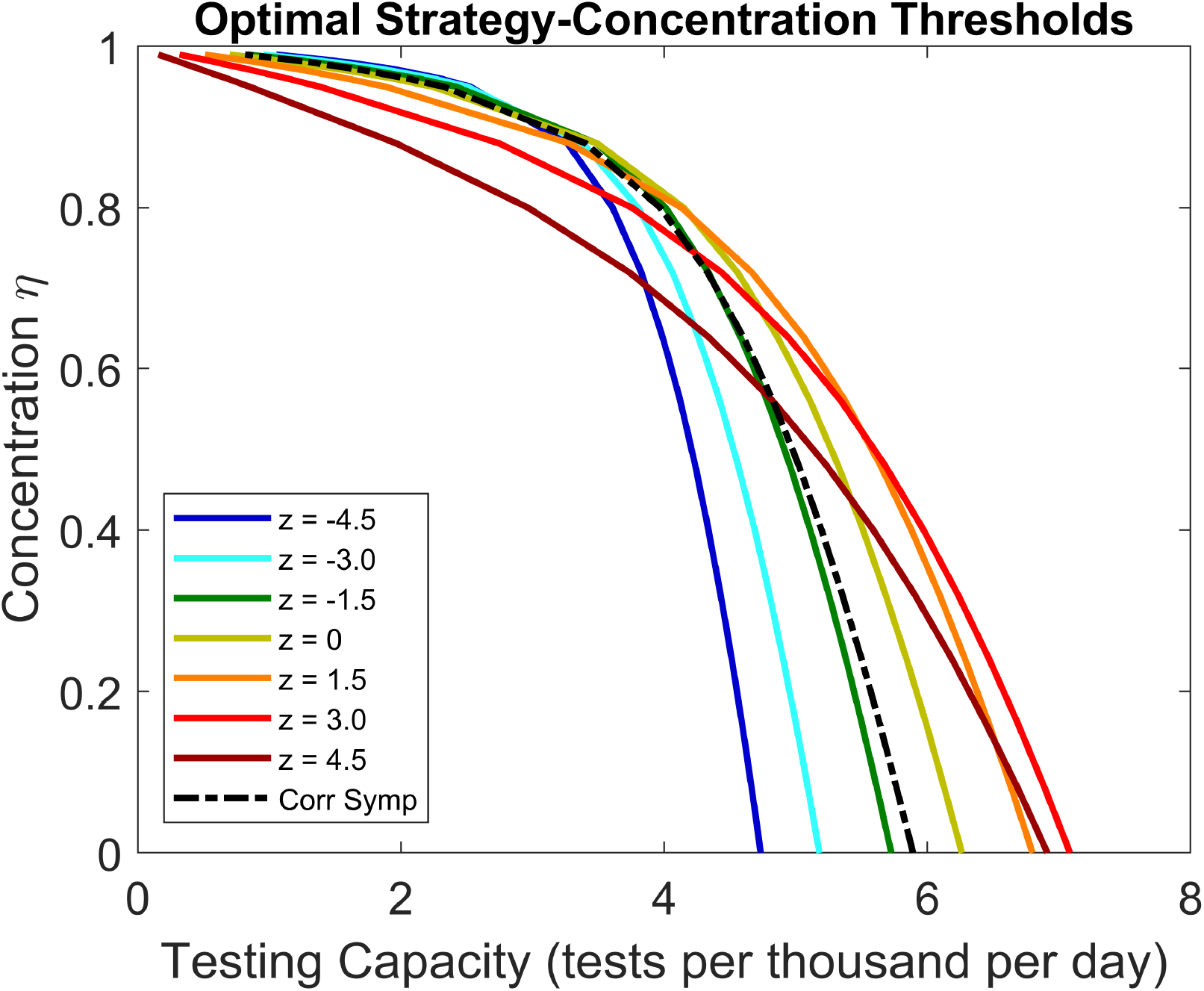
Strategy threshold testing capacities at various concentration levels *η* for the generic disease with 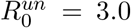. Curves represent (*C, η*) values where the strategy threshold capacity *C*^*th*^ occurs, assuming either incubation symptoms (colored curves) or correlated symptoms (black dashed curve) with gamma distributed *f*_*r*_ and *f*_*ε*_. Colors represents different off sets *z* between the mean incubation and mean latent period, with values in the legend given in units of days. At (*C, η*) points above or to the right of a given curve, optimal strategies are mixed clinical and non-clinical for that *z* value or symptom assumption. At points below or to the left, optimal strategies are clinical only. The *η* value for which a colored curve crosses the black curve represents a threshold *η*^*th*^ for which the role of non-clinical testing switches between enhanced and diminished compared to the correlated symptoms assumption.

From Figs. 10 and 12 (as well as the corresponding Figs. S12 and S13 in the Supplementary Material), it is clear that threshold concentrations *η*^*th*^ depend on both the incubation-latent offset *z* and the overall transmissibility as measured by 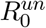, and may also depend on the mean latent period length. To visualize these influences, we find the thresholds *η*^*th*^ numerically for the *z* ≤ 0 diseases as well as the COVID-19 variants, assuming a spectrum of 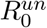 values for each model by changing the overall transmissibility. The results are plotted in Fig. 13 as curves in the 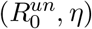 plane, where each curve represents *η*^*th*^ values for a different set of gamma distributed latent and incubation periods taken from Tables 2, 3, 4, and 6, assuming gamma distributions for *f*_*r*_. If a disease management official is designing a testing resource allocation strategy to control a disease for which they know the basic reproduction number and an estimate for their testing program’s *η* reproduction, they can locate the corresponding point in Fig. 13. If their point falls above the relevant incubation/latent period assumption curve, then due to presymptomatic transmission, optimal strategies will call for an increase in non-clinical testing resources relative to a comparable simpler disease for which infectiousness and symptoms always coincide. If their point falls below the relevant incubation/latent period assumption curve, optimal strategies will call for a reduction in non-clinical testing relative to a comparable simpler disease despite the presence of presymptomatic transmission. Generally, smaller 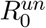 values require greater *η* to justify allocating extra resources to non-clinical testing, thus corroborating our previous observation that diseases with greater infectiousness require less precise non-clinical testing programs (i.e smaller *η*) to justify allocating additional resources to non-clinical testing due to presymptomatic transmission. We see further that larger *z* diseases and diseases with shorter latent periods (compare the omicron curve to the *z* = 1.50 curve) likewise require less precision to justify increasing resources to non-clinical testing. In other words, the factors of larger 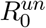, larger *z*, and smaller latent period expand the parameter space for which non-clinical testing is enhanced in optimal strategies due to presymptomatic transmission. Note that as 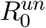 falls below 2.0, enhanced non-clinical testing is never justified. Thus, diseases with sufficiently low infectiousness do not warrant additional non-clinical testing resources, even if presymptomatic transmission may be significant.

**Figure 13.**
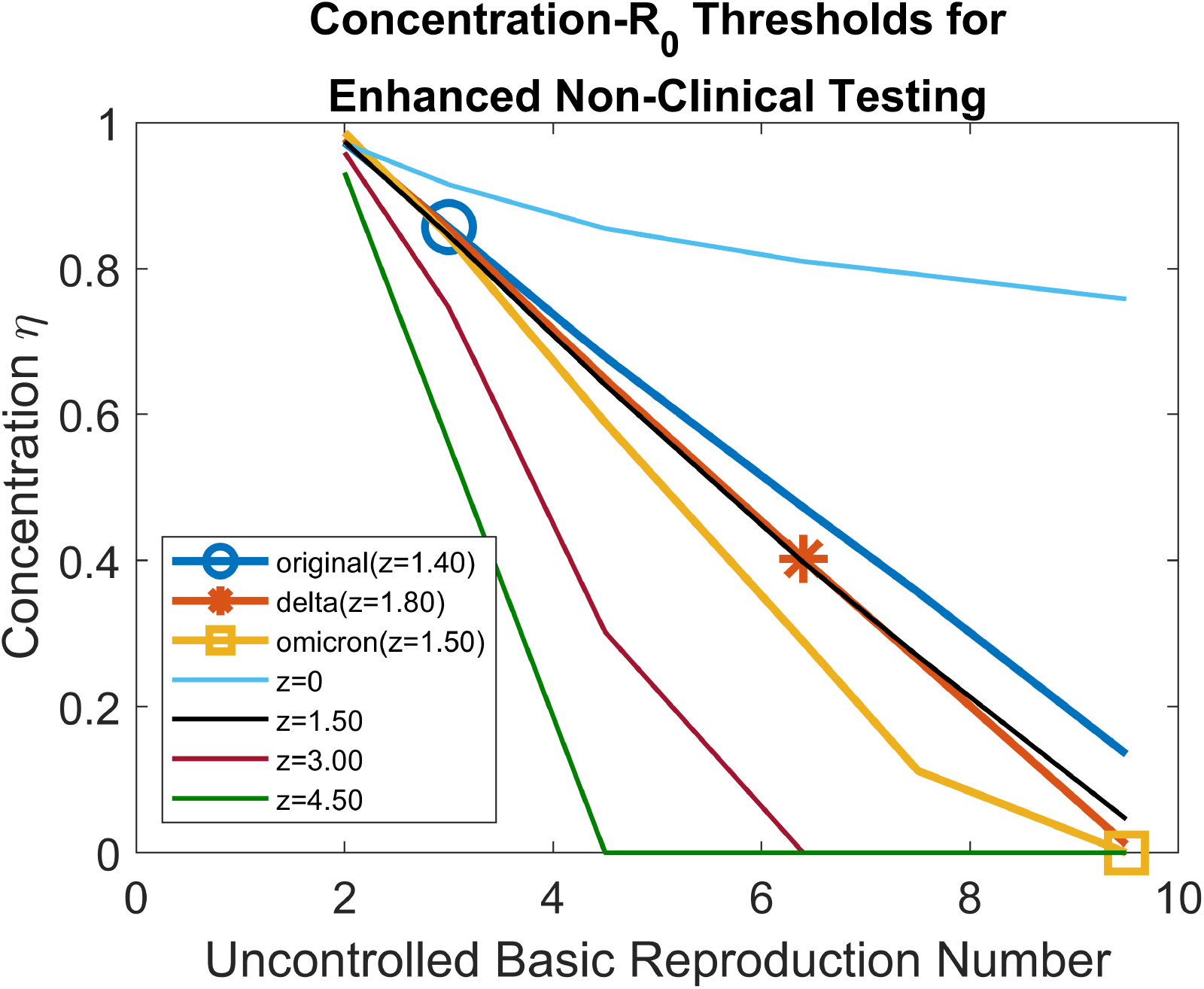
Curves depicting threshold concentrations *η*^*th*^ as a function of the uncontrolled basic reproduction number for gamma distributed latent and incubation period associated with the COVID-19 variants and our generic diseases, where *f*_*r*_ is assumed to be gamma distributed. Each curve represents a different latent/incubation period set that has a particular incubation-latent offset *z* denoted in the legend in units of days. The legend specifies the COVID variants along with their *z* offsets, and the generic disease indicated by only their *z* offsets. The actual 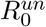 values for the COVID variants are indicated by makers on the plot. Points above a given curve represent the parameter space over which presymptomatic transmission justifies increasing resources to non-clinical testing for a disease with the corresponding latent and incubation periods. For points below a curve, presymptomatic transmission does not justify increasing non-clinical testing resources in optimal strategies.

## 4 Discussion

### 4.1 Curbing epidemics under resource limitations

The results of our work build a robust mathematical justification for the optimality of the testing strategies adopted by real-world disease management planners during the early phases of the COVID-19 pandemic. Namely, when testing resources are in short supply, they should be reserved for the most symptomatic and critically ill patients (6, 7, 8). Although these policies may have been adopted to prevent mortality, we have found that they are also good policies for suppressing an epidemic. More generally, our work shows that such policies are optimal for a broad range of disease characteristics and non-clinical testing program characteristics. Diseases with longer or shorter latent periods, stronger or weaker transmissibility, presymptomatic transmission or symptom onset before infectiousness or correlated symptom-infectiousness onset, and exponential or gamma period distributions all call for qualitatively similar policies as a function of testing capacity; optimal protocols call for clinical-only testing at testing capacities below a threshold *C*^*th*^, and call for mixed clinical and non-clinical strategies at greater testing capacities. Interestingly, in a previous work (18) where we analyzed optimally reducing the epidemic peak height (rather than total infection size) using the reduced and simplified ODE counterpart of our model here, we observed the same threshold behavior separating testing capacity regions between optimal clinical-only and mixed strategies. Thus, the threshold behavior appears to be a general feature of optimal allocation strategies under limited testing resources.

Figures 6, 7, and 11 depicting optimal total infection size for “realistic” and “random” non-clinical testing programs illuminate the feasibility or infeasibility of curbing a novel disease outbreak with a testing and quarantine program. Random testing (*η* = 0) is unlikely to be successful at the low testing capacities expected at the initial stages of a novel disease outbreak as production of newly developed tests falls well behind demand, even if the entire population is accessible and amenable to the limited number of tests available (*κ* = 1). However, significant disease suppression is possible at larger concentrations *η* even if the overall accessibility *κ* is small. This result stresses the importance of early implementation of effective contact tracing and public outreach programs encouraging individuals with suspected or probable recent transmission to seek testing, even if significant portions of the population are not open or accessible to such efforts. Importantly, this conclusion holds for the spectrum of model assumptions and disease characteristics analyzed in this paper.

### 4.2 Influence of model assumptions on controllability

Throughout this work, we have identified a number of disease characteristics and model assumptions that influence predictions of controllability. Model assumptions regarding latent period, incubation period, and recovery day distributions as well as incubation-latent offsets are critically important, as we have found that the exponential distribution and correlated symptom assumptions in the equivalent ODE model can severely overestimate controllability relative to the full IPDE model for the COVID-19 variants. The gamma distribution assumptions for *f*_*ε*_, *f*_*I*_, and *f*_*r*_ used in the full IPDE model are supported by epidemiological data and are far more realistic than ODE assumptions, so the reduced controllability exhibited by our model is a closer reflection of reality. Our model’s ability to decouple symptom onset from disease compartments plays a key role in its capacity to simultaneously incorporate realistic latent and incubation period distributions.

A major contributing factor to reduced controllability in the full IPDE model is the gamma period distribution for *f*_*r*_. This observation accords with previous models with identification + isolation/quarantine controls, where it was observed that an exponentially distributed infectious period can significantly overestimate controllability relative to a gamma distribution (27). In essence, quarantining is more effective under the exponential assumption due to its ability to significantly reduce the infectious period length of individuals in the long tail of the distribution (see Fig. 3). Likewise, under the exponential assumption, significantly larger numbers individuals recover naturally within the first few days of infectiousness onset in comparison to the gamma assumption, so delays in testing due to low capacity are irrelevant in these cases and are thus less detrimental to controllability. Conversely, for the latent period distribution, a gamma distribution assumption increases controllability relative to the exponential assumption (although to a lesser degree than that with which a gamma *f*_*r*_ reduces controllability) for reasons analogous to those of infectious period. Namely, under the exponential assumption, much larger numbers of individuals rapidly transition from exposed to infectious in comparison to the gamma distribution, so there is less time to locate and remove new infections before they begin generating new transmissions, and delays in identifying and quarantining infections due to supply limitations are thus more detrimental. On a more basic level, although gamma distributions for *f*_*ε*_ and *f*_*r*_ have no noticeable effect on total infection size under no control, they flatten/delay and sharpen/accelerate the uncontrolled epidemic peak, respectively, as compared to exponential distributions (see the Supplementary Material section S3). Intuitively, a disease that peaks earlier and to a greater degree on its on volition will be more difficult to control.

Symptom onset assumptions based on the age-of-infection can reduce or increase controllability depending on the incubation-latent offset *z*, as controllability generally decreases as *z* increases. Under correlated symptoms, active spreaders in the ES class always show symptoms and are thus always targeted with efficacious clinical testing. Under incubation symptoms, however, positive *z* values imply presymptomatic transmissions, meaning that some of the active spreaders can only be targeted with non-clinical testing which is subject to delays due to resources being wasted on uninfected individuals. These delays become more detrimental as the presymptomatic transmission window becomes larger, so controllability tends to decrease as *z* increases. For negative *z*, however, some individuals become symptomatic while still in the exposed class, so efficacious clinical testing can be used to remove infected individuals with minimal delay before they can begin generating new transmissions. This is impossible under correlated symptoms or positive *z* values. The opportunity for clinical testing to remove preinfectious individuals increases as *z* grows in the negative direction, and controllability consequently increases. Notably, the increase in controllability can be significant for a negative *z* disease compared to a positive *z* disease. This observation may explain why the 2003 SARS outbreak (negative *z* disease) was far easier to control than the COVID-19 pandemic (positive *z* disease) (54).

The remaining controllability reducing factors are straightforwardly explained. Namely, a more transmissible disease (i.e. one with a larger 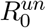) is inherently more difficult to control than a less transmissible disease, and the larger number of secondary infections generated by an individual can be detrimental to control when limited resources force delays in testing and quarantine. Likewise, shorter latent periods leave less time to locate infected individuals before they begin generating transmissions, so testing and quarantine delays become more costly. These factors together help explain why the omicron variant has been particularly challenging to control (49, 55). The large 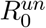 value is an oft emphasized contributing factor, but the short latent period relative to other variants can be just as important. Explicitly, comparing the generic disease (mean latent period 6 days) to the omicron variant (mean latent period 1.5 days) in Fig. 7 shows that omicron is roughly half as controllable despite both diseases having the same 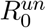.

### 4.3 Controllability, symptom onset, and the role of non-clinical testing

Our results across all model assumptions demonstrate a consistent clear relationship between non-clinical testing and controllability reducing factors unrelated to symptom onset assumptions: factors that reduce controllability also reduce the role of non-clinical testing in optimal strategies. We hypothesize that this relationship arises because wasted or inefficiently used resources are of greater detriment in a less controllable system. That is, when a disease is considered less controllable, failure to utilize a quantity of testing capacity to identify and isolate infected individuals will result in a greater number of additional infections in comparison to a more controllable disease. With non-clinical testing, the beneficial impact of identifying removing non-symptomatic infected individuals is counterbalanced by the negative impact of potentially wasting tests on uninfected individuals, and the negative impact becomes more prominent as the disease becomes less controllable. Thus, optimal strategies place more emphasis on clinical testing which is guaranteed to be utilized to identify and isolate an infected individual. However, for sufficiently large testing capacity, resources are plentiful enough such that wasting a test on an uninfected individual does not significantly delay or inhibit identification of infected individuals, so the role of non-clinical testing can be prominent in a less controllable disease, although still slightly diminished in comparison to a more controllable disease. Thus, the observed relationship between the role of non-clinical testing and controllability is closely tied to the resource limitation aspects of model.

In contrast to factors unrelated to symptom onset, the relationship symptom onset assumptions and non-clinical testing is unclear. One one hand, one might expect that for a negative *z* disease like COVID-19, realistically modeling the incubation-latent offset will increase the role of of non-clinical testing in comparison to a model making the simpler correlated symptom assumption due to the ability of non-clinical testing to capture presymptomatic spreaders. On the other hand, since the latent incubation offset decreases controllability for a negative *z* disease, one might expect the role of non-clinical testing to be diminished due to the reasoning given in the preceding paragraph. We found that neither of these hypotheses are generally correct for guiding optimal strategies based on the results of simple ODE models. Instead, we found that age-of-infection modeling, and in particular, the details of the temporal offset between the incubation and latent periods, was critical to understanding disease progression, controllability, and optimal strategies for resource allocation. Our results show that optimal policies for resource allocation depend on a complicated (often non-intuitive) interplay between incubation-latent offset, disease strength, and other characteristics like latent period length (Figs. 11, 12, 13 and the corresponding Figs. S10, S11, S12, S13 in the Supplementary Material). This is a critical observation because real-world public policies and attitudes towards a novel diseases are typically based on what has been successful for previous diseases combined with basic intuition for the broad differences between the novel disease and previous diseases. For example, 2003 SARS has no presymptomatic transmission while COVID-19 has significant presymptomatic transmission, so one might intuitively expect that a successful strategy for controlling 2003 SARS should be adapted for COVID-19 by allocating more resources to non-clinical testing to capture presymptomatic spreaders, but this is not at all the case according to our results. Our work shows that this intuition-based approach to public policy can fail, and thus highlights the importance of mathematical modeling in helping to guide disease management. In particular, it is important to establish modeling frameworks where different diseases can be examined comparatively under the same lens to more closely mimic the way diseases are compared in the real world. Utilizing the age-of-infection to decouple symptom status from specific model compartments is the crucial element for allowing such comparisons in our model. Future models may build upon these ideas to incorporate not only the age-of-infection, but also the age-of-infectiousness to model additional realistic features like partial correlations between incubation, latent, and infectious period lengths as well as time-varying infectivity levels dependent on the time since infectiousness or symptom onset.

## Supporting information

Supplementary Material Sections

## Data Availability

All data produced in the present study are available upon reasonable request to the authors.

## Acknowledgments

This work was funded by the Center of Advanced Systems Understanding (CASUS) which is financed by Germanys Federal Ministry of Education and Research (BMBF) and by the Saxon Ministry for Science, Culture and Tourism (SMWK) with tax funds on the basis of the budget approved by the Saxon State Parliament.

