## Supplementary Material Sections for "The relationship between controllability, optimal testing resource allocation, and incubation-latent period mismatch as revealed by COVID-19"

#### S1 Basic reproduction number derivation

We derive an expression the basic reproduction number $R_0$ for our model using the next-generation operator method for compartmental models (1, 2). Suppose at some arbitrary time $t = 0$ a small number of newly infected (i.e. age of infection $x = 0$) individuals $\mathbf{g}_0$ is introduced into an otherwise disease free system, where the components of the vector $\mathbf{g}_0$ denote the following infection classes:

$$\mathbf{g}_0 = \begin{pmatrix} E_A \\ E_Y \\ A \\ Y \end{pmatrix}. \tag{s1}$$

Let $\mathbf{g}(x)$ denote the infection status of those individuals who are still infected after $x$ infection days. Assuming that the number of new infections generated by these individuals remains small, we linearize our model equations (Eq. (7) in the main text) about the disease-free state to find the following ODE for the time evolution of $\mathbf{g}(x)$:

$$\frac{d\mathbf{g}(x)}{dx} = -V(x)\mathbf{g}(x), \ \ \mathbf{g}(0) = \mathbf{g}_0, \tag{s2}$$

where $V(x)$ denotes the following $4 \times 4$ matrix:

$$V(x) = \begin{pmatrix} V_{11}(x) & 0 & 0 & 0 \\ 0 & V_{22}(x) & 0 & 0 \\ V_{31}(x) & 0 & V_{33}(x) & 0 \\ 0 & V_{42}(x) & 0 & V_{44}(x) \end{pmatrix}, \tag{s3}$$

where

$$\begin{aligned} V_{11}(x) &= \varepsilon(x) + \tau_{N_0}^{-1}\kappa \\ V_{22}(x) &= \varepsilon(x) + \tau_{N_0}^{-1}\kappa(1 - P_e(x)) + \tau_{C_0}^{-1}P_e(x) \\ V_{33}(x) &= r(x) + \tau_{N_0}^{-1} \\ V_{44}(x) &= r(x) + \tau_{N_0}^{-1}\kappa(1 - P_y(x)) + \tau_{C_0}^{-1}P_y(x) \\ V_{31}(x) &= -\varepsilon(x) \\ V_{42}(x) &= -\varepsilon(x), \end{aligned} \tag{s4}$$

and where $\tau_{N_0}^{-1}$ and $\tau_{C_0}^{-1}$ are the non-clinical and clinical testing rates, respectively, evaluated at the disease free equilibrium:

$$\tau_{N_0}^{-1} = \begin{cases} 0, & C = 0 \text{ or } \rho = 0 \\ \left[\tau + \kappa \frac{1-\eta}{\rho C}\right]^{-1}, & \text{otherwise}, \end{cases} \tag{s5}$$

$$\tau_{C_0}^{-1} = \begin{cases} 0, & C = 0 \text{ or } \rho = 1 \\ \tau^{-1}, & \text{otherwise}. \end{cases} \tag{s6}$$

The component $-V_{ij}(x)$ is the transition rate from infection class $i$ to infection class $j$ for $i \neq j$, while the diagonal component $V_{ii}(x)$ is the total rate of transition out of infection class $i$, assuming an otherwise disease free system. The solution to Eq. (s2) is written as the following:

$$\mathbf{g}(x) = M(x)\mathbf{g}_0, \tag{s7}$$

where $M(x)$ is the solution to the following matrix differential equation:

$$\frac{dM(x)}{dx} = -V(x)M(x), \; M(0) = \mathcal{I}, \tag{s8}$$

where $\mathcal{I}$ denotes the $4 \times 4$ identity matrix. The solution to Eq. (s8) is given by the following:

$$M(x) = \begin{pmatrix} M_{11}(x) & 0 & 0 & 0 & \\ 0 & M_{22}(x) & 0 & 0 & \\ M_{31}(x) & 0 & M_{33}(x) & 0 & \\ 0 & M_{42}(x) & 0 & 0 & M_{44}(x) \end{pmatrix}, \tag{s9}$$

where

$$\begin{aligned}
M_{11}(x) &= \exp\left[-\int_0^x ds\left(\varepsilon(s) + \tau_{N_0}^{-1}\kappa\right)\right] \qquad \text{(s10)}\\
M_{22}(x) &= \exp\left[-\int_0^x ds\left(\varepsilon(s) + \tau_{N_0}^{-1}\kappa\big(1 - P_e(s)\big) + \tau_{C_0}^{-1}P_e(s)\right)\right]\\
M_{33}(x) &= \exp\left[-\int_0^x ds\left(r(s) + \tau_{N_0}^{-1}\kappa\right)\right]\\
M_{44}(x) &= \exp\left[-\int_0^x ds\left(r(s) + \tau_{N_0}^{-1}\kappa\big(1 - P_y(s)\big) + \tau_{C_0}^{-1}P_y(s)\right)\right]\\
M_{31}(x) &= \int_0^x ds\,\varepsilon(s)\exp\left[-\int_0^s ds'\left(\varepsilon(s') + \tau_{N_0}^{-1}\kappa\right) - \int_s^x ds'\left(r(s') + \tau_{N_0}^{-1}\kappa\right)\right]\\
M_{42}(x) &= \int_0^x ds\,\varepsilon(s)\exp\Bigg[-\int_0^s ds'\left(\varepsilon(s') + \tau_{N_0}^{-1}\kappa(1 - P_e(s')) + \tau_{C_0}^{-1}P_e(s')\right)\\
&\qquad - \int_s^x ds'\left(r(s') + \tau_{N_0}^{-1}\kappa(1 - P_y(s')) + \tau_{C_0}^{-1}P_y(s')\right)\Bigg].
\end{aligned}$$

The number of new infections generated by $\mathbf{g}(x)$ in the otherwise disease-free system is given by the following matrix integral :

$$\int_0^\infty dx\, F\mathbf{g}(x) = \int_0^\infty dx\, FM(x)\mathbf{g}_0, \tag{s11}$$

where $F$ denotes the $4 \times 4$ infection rate matrix:

$$F = \begin{pmatrix} 0 & 0 & f_A \lambda_A \beta & f_A \lambda_Y \beta \\ 0 & 0 & f_Y \lambda_A \beta & f_Y \lambda_Y \beta \\ 0 & 0 & 0 & 0 \\ 0 & 0 & 0 & 0 \end{pmatrix}. \tag{s12}$$

The component $F_{ij}$ represents the rate of new infections generated in infection class $i$ by an individual in infection class $j$ in an otherwise disease free system. Expressions for these components are found by linearizing $e_A(t,0), e_Y(t,0), a(t,0)$, and $y(t,0)$ in Eq. (7) of the main text about the disease-free equilibrium. The matrix in Eq. (s11) defines next generation matrix $\mathcal{N}$ (1, 2):

$$\mathcal{N} = \int_0^\infty dx\, F M(x). \tag{s13}$$

The basic reproduction number $R_0$ is defined as the spectral radius of $\mathcal{N}$, i.e. the largest solution to the eigenvalue problem $\mathcal{N}\mathbf{g}_0 = R_0 \mathbf{g}_0$ which is guaranteed to be real and non-negative (1, 2). The following analytic expression for $R_0$ is obtained by finding the largest positive solution to the determinant equation $\det(\mathcal{N} - R_0 \mathcal{I}) = 0$:

$$R_0 = \int_0^\infty dx \int_0^x ds\, \alpha_A(x,s) + \int_0^\infty dx \int_0^x ds\, \alpha_Y(x,s) \tag{s14}$$

where we define the following notation:

$$\begin{aligned} \alpha_A(x,s) &= f_A \lambda_A \beta\, \varepsilon(s) \exp\left[ -\int_0^s ds' \left( \varepsilon(s') + \tau_{N_0}^{-1} \kappa \right) - \int_s^x ds' \left( r(s') + \tau_{N_0}^{-1} \kappa \right) \right] \\ \alpha_Y(x,s) &= f_Y \lambda_Y \beta\, \varepsilon(s) \exp\left[ -\int_0^s ds' \left( \varepsilon(s') + \tau_{N_0}^{-1} \kappa (1 - P_e(s')) + \tau_{C_0}^{-1} P_e(s') \right) \right. \\ &\qquad \left. - \int_s^x ds' \left( r(s') + \tau_{N_0}^{-1} \kappa (1 - P_y(s')) + \tau_{C_0}^{-1} P_y(s') \right) \right] \end{aligned} \tag{s15}$$

The $\alpha_A$ double integral Eq. (s14) is the contribution to $R_0$ from never symptomatic infections while the $\alpha_Y$ double integral is the contribution from eventually symptomatic infections.

#### S2 Basic reproduction number analysis

Considered as a function of the resource allocation strategy parameter $\rho$, $R_0(\rho)$ is differentiable for $\rho \in [0,1)$ and is discontinuous at $\rho = 1$ whenever $C > 0$ such that $R_0(1) > \lim_{\rho\to 1^-} R_0(\rho)$. Taking the derivative of $R_0(\rho)$ with respect to $\rho$ yields the following:

$$
\begin{aligned}
\frac{dR_0(\rho)}{d\rho} &= -\int_0^\infty dx \int_0^s ds\, \alpha_A(x,s) \int_0^x ds' \kappa \frac{d\tau_{N_0}^{-1}}{d\rho} \\
&\quad -\int_0^\infty dx \int_0^s ds\, \alpha_Y(x,s) \left[ \int_0^s ds' \kappa \frac{d\tau_{N_0}^{-1}}{d\rho} (1 - P_e(s') + \int_s^x ds' \kappa \frac{d\tau_{N_0}^{-1}}{d\rho} (1 - P_y(s')) \right] \\
&= -[\kappa \tau_{N_0}^{-1}]^2 \frac{1-\eta}{\rho^2 C} \int_0^\infty dx \int_0^s ds\, \alpha_A(x,s) x \\
&\quad -[\kappa \tau_{N_0}^{-1}]^2 \frac{1-\eta}{\rho^2 C} \int_0^\infty dx \int_0^s ds\, \alpha_Y(x,s) \left[ \int_0^s ds' (1 - P_e(s') + \int_s^x ds' (1 - P_y(s')) \right].
\end{aligned}
\tag{s16}
$$

The integrands in the above equation are positive, so $\frac{dR_0(\rho)}{d\rho} \leq 0$ on $\rho \in [0,1)$, and $\lim_{\rho\to 1^-} R_0(\rho)$ is therefore the greatest lower bound for $R_0(\rho)$ on $\rho \in [0,1]$. The expression for $\lim_{\rho\to 1^-} R_0(\rho)$ is obtained by substituting the following limits for the testing rates into Eq. (s14):

$$
\lim_{\rho\to 1^-} \tau_{N_0}^{-1} = \begin{cases} 0, & C = 0 \\ \left[\tau + \kappa \frac{1-\eta}{C}\right]^{-1}, & C > 0, \end{cases}
\tag{s17}
$$

$$
\lim_{\rho\to 1^-} \tau_{C_0}^{-1} = \begin{cases} 0, & C = 0 \\ \tau^{-1}, & C > 0. \end{cases}
\tag{s18}
$$

Under the ODE assumptions for our model (exponential distributions and correlated symptoms), we have $\varepsilon(x) = \varepsilon$, $r(x) = r$, $P_e(x) = 0$, and $P_y(x) = 1$, and $R_0$ simplifies to the following expression:

$$
R_0 \underset{ODE}{=} \frac{\varepsilon}{\varepsilon + \kappa \tau_{N_0}^{-1}} \frac{f_A \beta \lambda_A}{r + \kappa \tau_{N_0}^{-1}} + \frac{\varepsilon}{\varepsilon + \kappa \tau_{N_0}^{-1}} \frac{f_Y \beta \lambda_Y}{r + \tau_{C_0}^{-1}}.
\tag{s19}
$$

For the uncontrolled system, where $C = 0$ such that $\tau_{N_0}^{-1} = \tau_{C_0}^{-1} = 0$, we find the following uncontrolled basic reproduction number $R_0^{un}$:

$$\begin{aligned} R_0^{un} &= \int_0^\infty dx \int_0^x ds \left(f_A\beta\lambda_A + f_Y\beta\lambda_Y\right)\varepsilon(s)\exp\left[-\int_0^s ds'\,\varepsilon(s') - \int_s^x ds'\,r(s')\right] \\ &\underset{ODE}{=} \frac{f_A\beta\lambda_A}{r} + \frac{f_Y\beta\lambda_Y}{r}. \end{aligned} \tag{s20}$$

The ODE expression for $R_0^{un}$ is equivalent to that of Eq. (12) in the main text. Values of $R_0^{un}$ for the COVID variants and various distribution assumptions are given in Table S1. Here, we fix $R_0^{un}$ under exponential distribution assumptions on both $f_\varepsilon$ and $f_r$ to values found in the literature by setting values for the transmissibilities $\lambda_A$ and $\lambda_Y = 2\lambda_A$. The same transmissibilities are used for non-exponential distribution assumptions, so changes in $R_0^{un}$ relative to the both exponential $f_\varepsilon$ and $f_r$ case represent changes due solely to the use of gamma distributions. We find that distribution assumptions have a very small impact on $R_0^{un}$.

| $R_0^{un}$ values | $f_\varepsilon$ Gamma Distribution | $f_\varepsilon$ Exponential Distribution |
|---|---|---|
| $f_r$ Gamma Distribution | Original: 3.004<br>Delta: 6.402<br>Omicron: 9.500 | Original: 3.098<br>Delta: 6.473<br>Omicron: 9.501 |
| $f_r$ Exponential Distribution | Original: 3.000<br>Delta: 6.400<br>Omicron: 9.500 | Original: 3.000<br>Delta: 6.400<br>Omicron: 9.500 |

Table S1: Uncontrolled basic reproduction number $R_0^{un}$ for COVID variants assuming either exponential or gamma distributions for $f_\varepsilon$ and $f_r$ (symptom onset assumptions are irrelevant in the uncontrolled system).

#### S3 Uncontrolled COVID variant infection curves

In Fig. S2, we plot outbreak curves under the uncontrolled disease dynamics for original, delta, and omicron COVID variants, assuming different combinations of exponential and gamma distributions for $f_r$ and $f_\varepsilon$ differentiated by curve color and dashing style as indicated in Fig. S1 (correlated versus incubation symptoms assumptions have no impact on the disease dynamics under no control). Although the results of the main text show that distribution assumptions have no noticeable impact on the total number of infections generated under the uncontrolled disease dynamics, Fig. S2 shows noticeable effects on the epidemic peak height and peak time. A gamma distributed $f_r$ tends to expedite peak times and significantly increase the peak heights. Conversely, a gamma distributed $f_\varepsilon$ tends to delay peak times and, when $f_r$ is gamma distributed, can marginally reduce peak heights. We hypothesize that distribution assumptions on $f_\varepsilon$ noticeably influence peak heights only for a gamma distributed $f_r$ because the infectious period distribution $f_{inf}$ is independent of $f_\varepsilon$ when $f_r$ is exponential (see Eq. (10) of the main text).

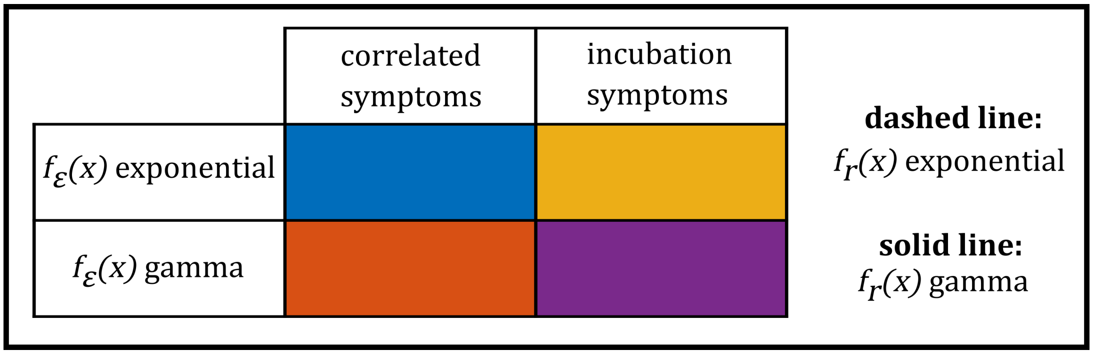


Figure S1: Legend for interpreting curve color and style in Figs. S2, S3, S4, S5, S6, S7, S9a, and S9b. Different colors represent different combinations of exponential and gamma distribution assumptions for $f_\varepsilon$ along with different assumptions for correlated versus incubation symptom onset as indicated by the colored table. Dashed lines indicate the exponential distribution assumption for $f_r$ while solid lines indicate the gamma distribution assumption. The model with all IPDE elements is given by a solid purple curve while the ODE equivalent model is given by a dashed blue curve.

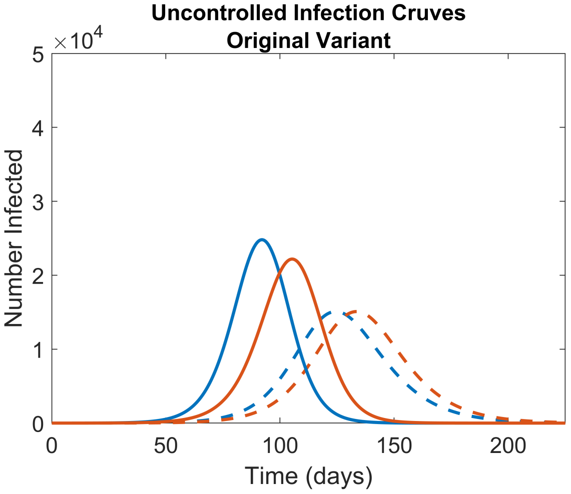


(a)

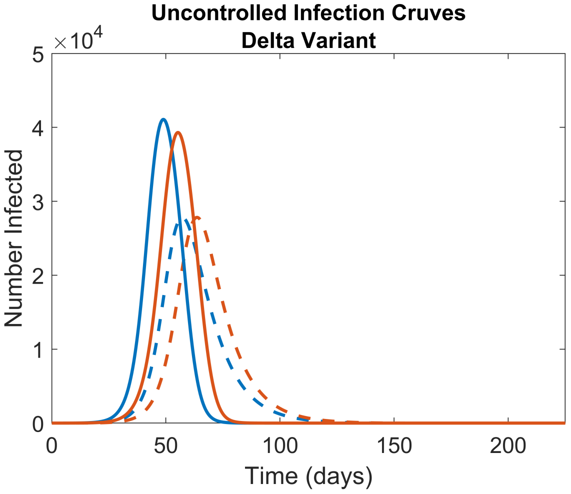


(b)

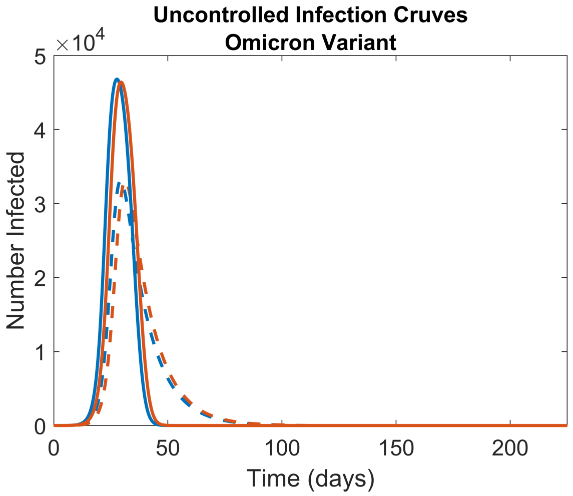


(c)

Figure S2: Uncontrolled disease dynamics for the 3 COVID variants assuming either exponential or gamma distributions for $f_r$ and $f_\varepsilon$. Curve color and solid versus dashed lines represent different distribution assumptions as indicated in Fig. S1. We plot curves only for the correlated symptoms (rather than incubation symptoms) because symptom onset assumptions have no impact on the uncontrolled disease dynamics.

### S4 Alternative measures of controllability

In the main text, we use the amount of testing capacity required to reduce total infections to a given level as a measure of controllability. Here, we present two alternative notions of controllability utilizing the basic reproduction number. These alternatives manifest the same same controllability reducing factors as listed in Table 7 of the main text, thus showing that our main text results and conclusions on controllability are quite general.

First, we consider the minimum testing capacity required to reduce $R_0$ to 1 for given set of model assumptions and testing parameters. A smaller minimum testing capacity indicates that disease outbreaks and be effectively suppressed with fewer resources and thus represents a greater degree of controllability. Because $\lim_{\rho \to 1^-} R_0(\rho)$ (henceforth denoted $R_0(1-)$) is the greatest lower bound for $R_0$ under a given set of distribution assumptions, testing parameters, and model parameters, the minimum testing capacity required to reduce $R_0$ to 1 is equivalent to the testing capacity required to reduce $R_0(1-)$ to 1. We find this minimal testing capacity numerically under a variety of model assumptions and present the results in Figs. S3, S4, S5, S6, S7, and S8. These plots show that factors which reduce controllability in the sense of raising the minimum testing capacity needed to reduce $R_0$ to 1 are the same factors which reduce controllability in the sense used in the main text.

The ability to reduce $R_0$ to 1 is not guaranteed in our model. Specifically, for any set of that model parameters and distribution assumptions, there may exist a critical $\kappa$ value below which $R_0(1-)$ is always greater than 1 even when unlimited resources are available (i.e $C \to \infty$). This critical $\kappa$ provides an additional alternative measure of controllability, where larger critical $\kappa$ indicates decreased ranges of parameter and model assumptions over which disease outbreaks can be suppressed. We calculate these critical $\kappa$ values for the COVID variants and generic disease under a variety of model assumptions and plot our results in Fig. S9. We find that factors that reduce controllability in the sense of raising the critical $\kappa$ are the same factors that reduce controllability in the sense used in the main text.

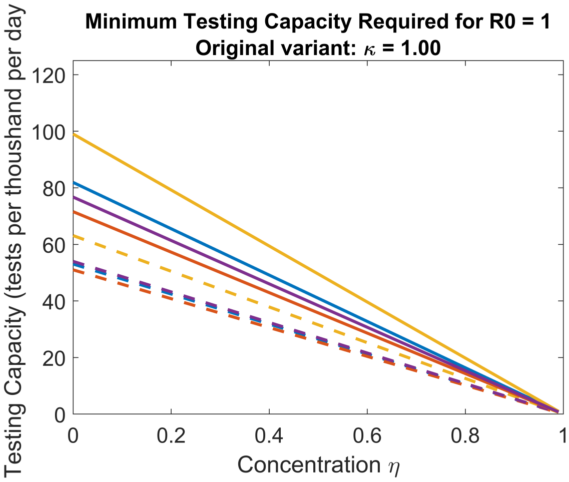


(a) Original Variant: Variable Distribution and Symptom Assumptions

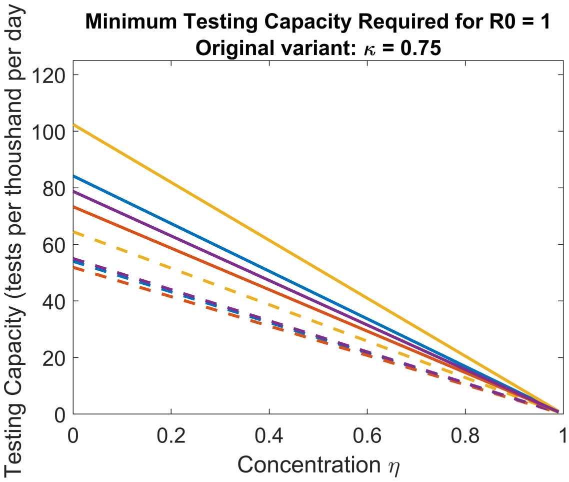


(b) Original Variant: Variable Distribution and Symptom Assumptions

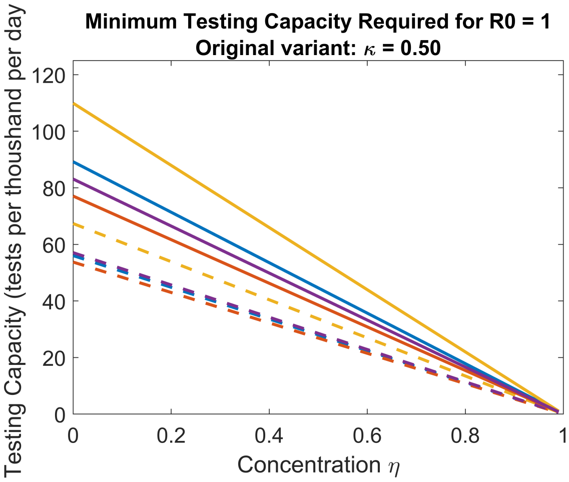


(c) Original Variant: Variable Distribution and Symptom Assumptions

Figure S3: Minimum testing capacities needed to reduce $R_0$ to 1 as a function of concentration $\eta$ for the original COVID variant, assuming a variety of distribution assumptions and accessibility levels $\kappa$. The meaning of curve color and dashed versus solid curves is given in Fig. S1.

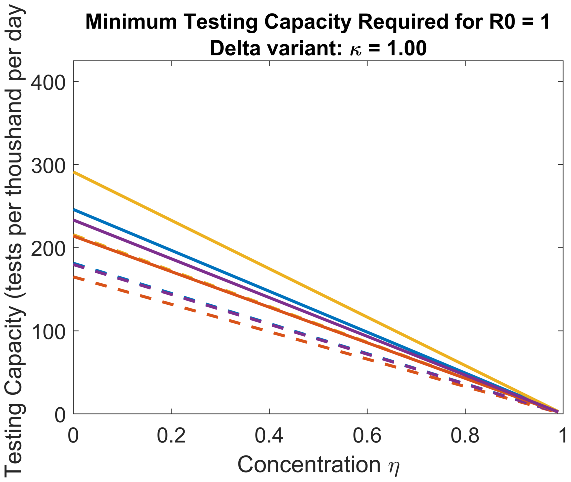


(a) Delta Variant: Variable Distribution and Symptom Assumptions

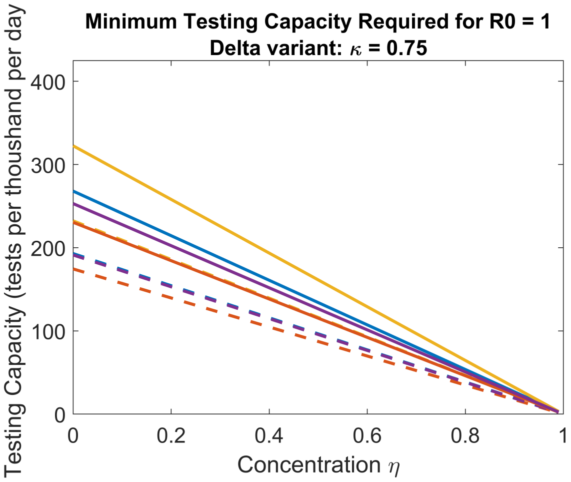


(b) Delta Variant: Variable Distribution and Symptom Assumptions

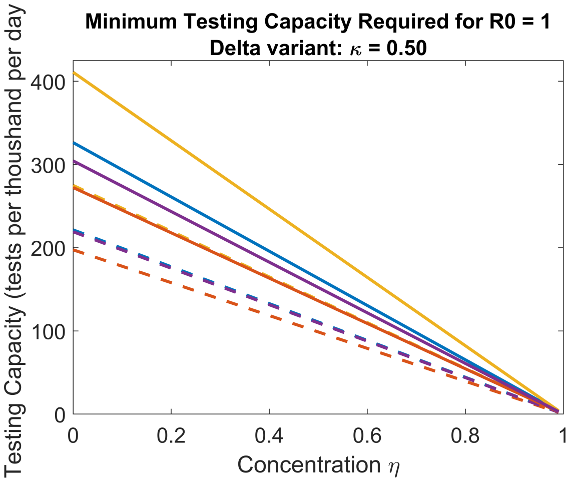


(c) Delta Variant: Variable Distribution and Symptom Assumptions

Figure S4: Minimum testing capacities needed to reduce $R_0$ to 1 as a function of concentration $\eta$ for the delta COVID variant, assuming a variety of distribution assumptions and accessibility levels $\kappa$. The meaning of curve color and dashed versus solid curves is given in Fig. S1.

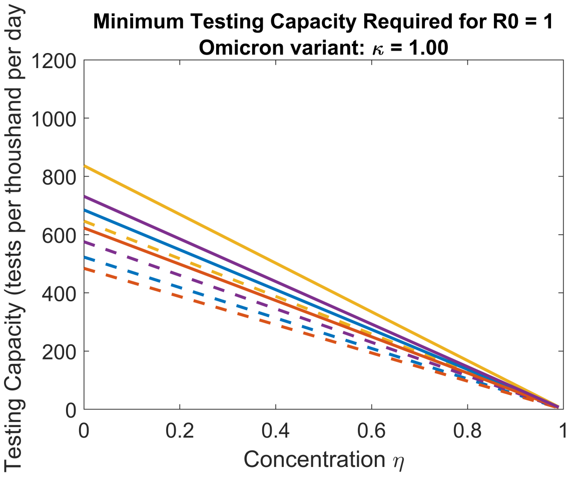


(a) Omicron Variant: Variable Distribution and Symptom Assumptions

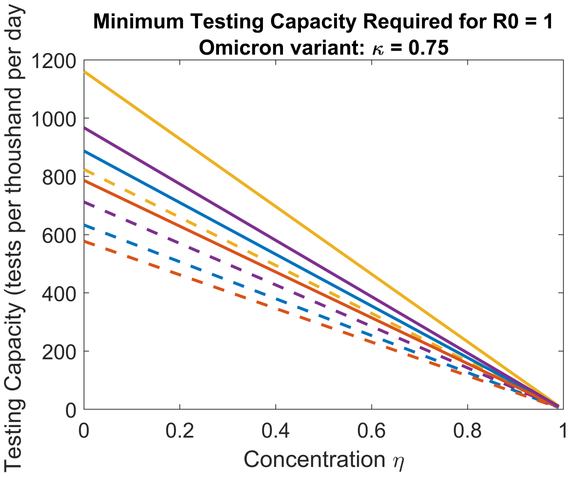


(b) Omicron Variant: Variable Distribution and Symptom Assumptions

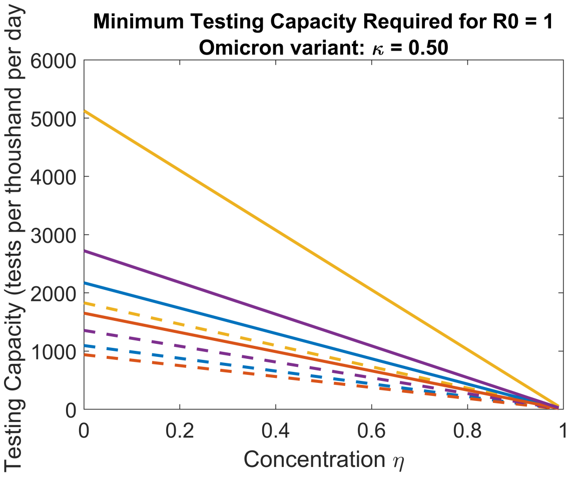


(c) Omicron Variant: Variable Distribution and Symptom Assumptions

Figure S5: Minimum testing capacities needed to reduce $R_0$ to 1 as a function of concentration $\eta$ for the Omicron COVID variant, assuming a variety of distribution assumptions and accessibility levels $\kappa$. Note the change in scale in Fig. S5c. The meaning of curve color and dashed versus solid curves is given in Fig. S1.

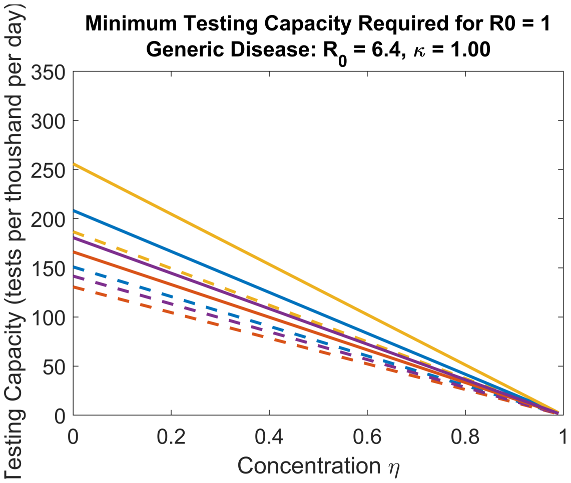


(a) Generic Disease $R_0^{un} = 6.4$, $z = 1.5$ days: Variable Distribution and Symptom Assumptions

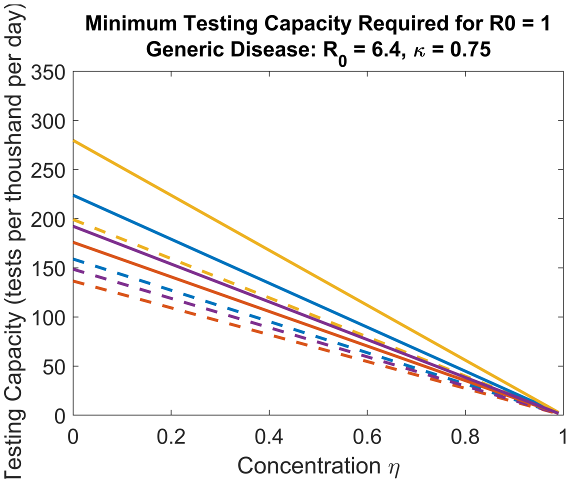


(b) Generic Disease $R_0^{un} = 6.4$, $z = 1.5$ days: Variable Distribution and Symptom Assumptions

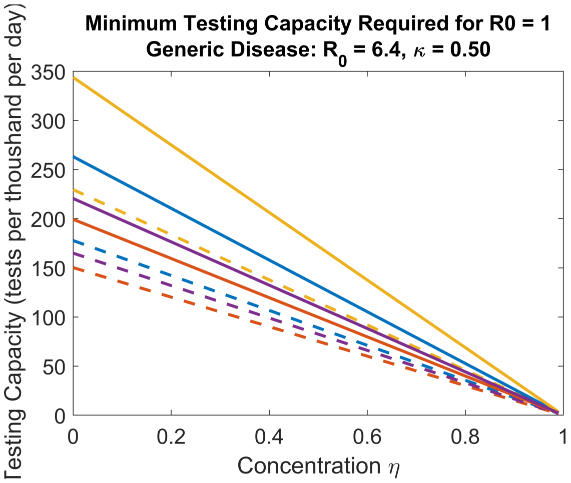


(c) Generic Disease $R_0^{un} = 6.4$, $z = 1.5$ days: Variable Distribution and Symptom Assumptions

Figure S6: Minimum testing capacities needed to reduce $R_0$ to 1 as a function of concentration $\eta$ for the generic disease with $R_0^{un} = 6.4$ and $z = 1.5$ days, assuming a variety of distribution assumptions and accessibility levels $\kappa$. The meaning of curve color and dashed versus solid curves is given in Fig. S1.

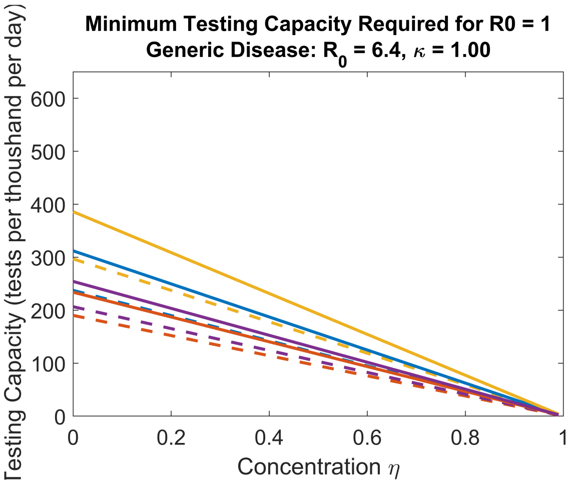


(a) Generic Disease $R_0^{un} = 9.5$, $z = 1.5$ days: Variable Distribution and Symptom Assumptions

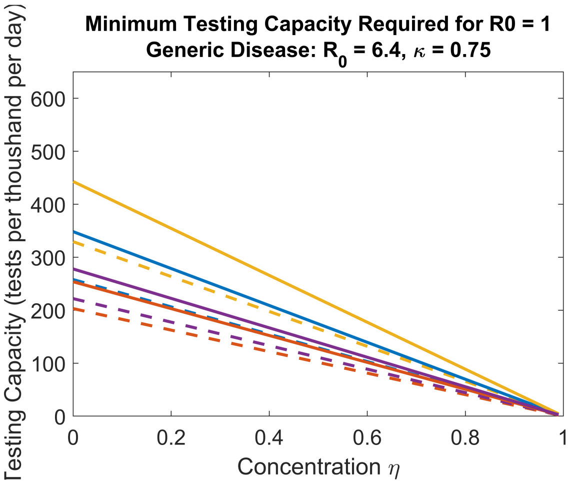


(b) Generic Disease $R_0^{un} = 9.5$, $z = 1.5$ days: Variable Distribution and Symptom Assumptions

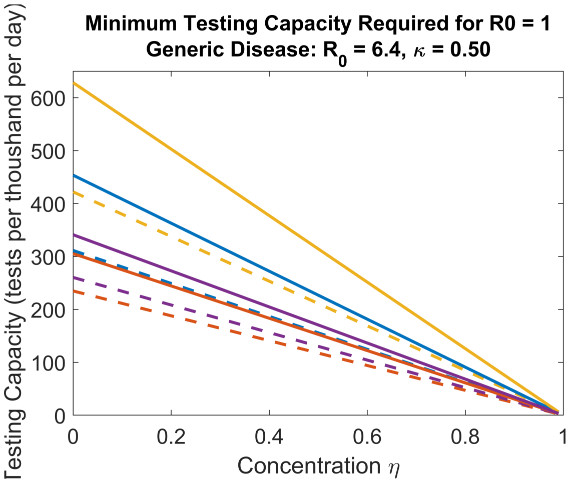


(c) Generic Disease $R_0^{un} = 9.5$, $z = 1.5$ days: Variable Distribution and Symptom Assumptions

Figure S7: Minimum testing capacities needed to reduce $R_0$ to 1 as a function of concentration $\eta$ for the generic disease with $R_0^{un} = 9.5$ and $z = 1.5$ days, assuming a variety of distribution assumptions and accessibility levels $\kappa$. The meaning of curve color and dashed versus solid curves is given in Fig. S1.

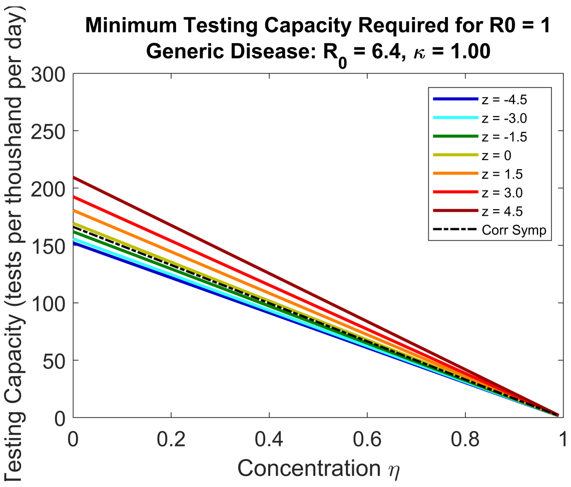


(a) Generic Disease $R_0^{un} = 6.4$: Gamma Distribution Assumptions and Variable Incubation-Latent Offsets

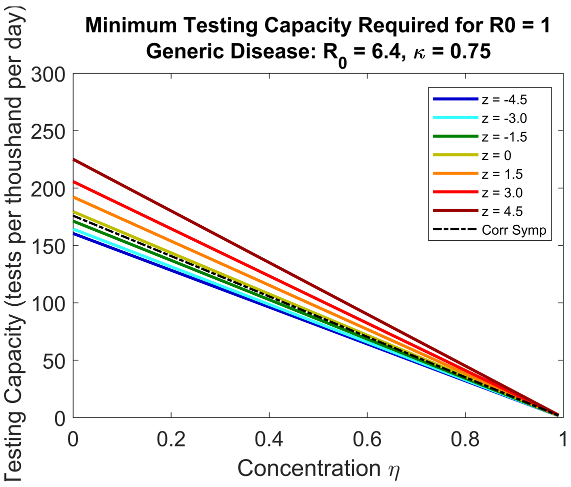


(b) Generic Disease $R_0^{un} = 6.4$: Gamma Distribution Assumptions and Variable Incubation-Latent Offsets

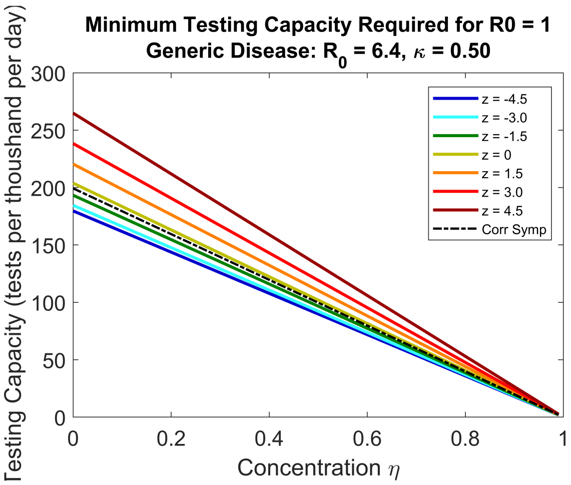


(c) Generic Disease $R_0^{un} = 6.4$: Gamma Distribution Assumptions and Variable Incubation-Latent Offsets

Figure S8: Minimum testing capacities needed to reduce $R_0$ to 1 as a function of concentration $\eta$ for the generic disease with $R_0^{un} = 6.4$, assuming gamma distributions for $f_\varepsilon$ and $f_r$ and various concentration $\kappa$ values. Curve colors represent different offsets $z = \langle f_I \rangle - \langle f_\varepsilon \rangle$ between mean incubation and latent periods as indicated by the legends (measured in units of days). Black dashed curves represent the correlated symptoms assumption where $z$ values are irrelevant.

### S5 Additional generic disease plots

The following plots are analogous to the generic disease optimal infection size and optimal strategy plots in Figs. (11) and (12) of the main text, but with greater disease strengths $R_0^{un} = 6.4$ and $R_0^{un} = 9.5$.

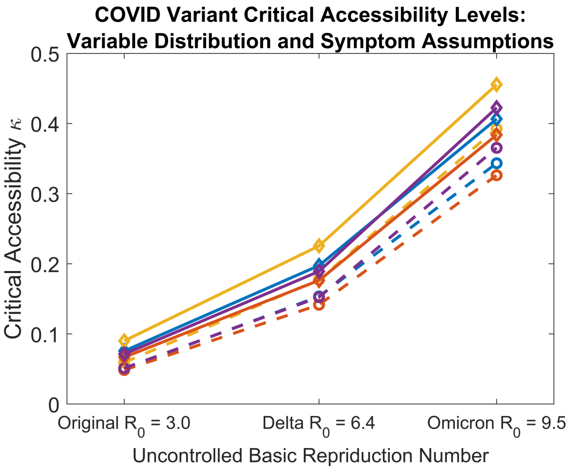


(a) COVID-19 Variants: Variable Distribution and Symptom Assumptions

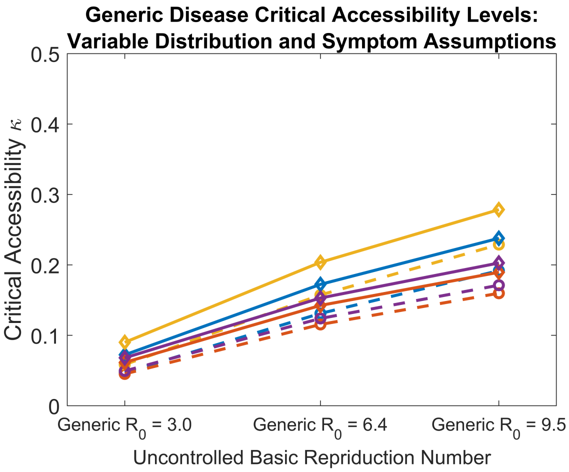


(b) Generic Disease $z = 1.5$ days: Variable Distribution and Symptom Assumptions

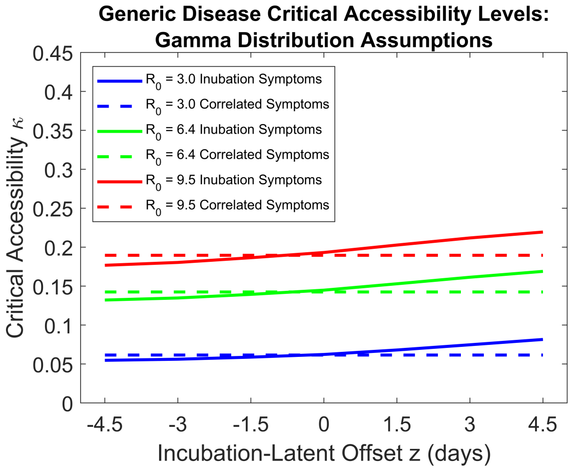


(c) Generic Disease: Gamma Distribution Assumptions and Variable $z$

Figure S9: Critical accessibility levels $\kappa$ below which $R_0$ can not be reduced to 1. Figures S9a and S9b plot critical accessibility as a function of uncontrolled $R_0$ for the COVID variants and the comparable generic disease with incubation-latent offset $z = 1.5$ days under various distribution and symptom onset assumptions, where the meaning of curve color and dashed versus solid curves is given in Fig. S1. Figure S9c plots critical accessibility for the generic disease as a function of incubation-latent offset, where differnt curve colors represent different uncontrolled $R_0$ values.

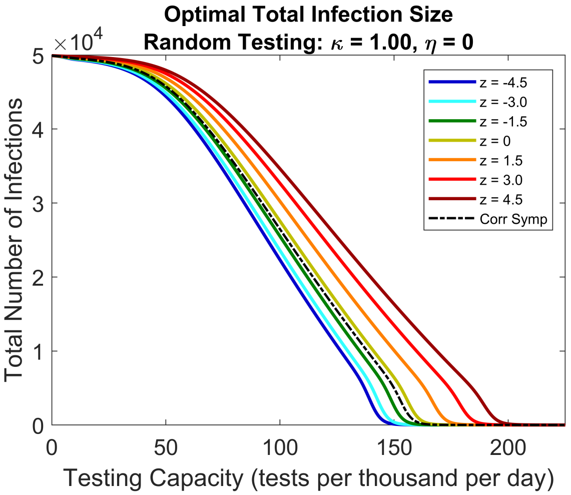


(a) Generic Disease $R_0^{un} = 6.4$: Gamma Distribution Assumptions and Variable Incubation-Latent Offsets

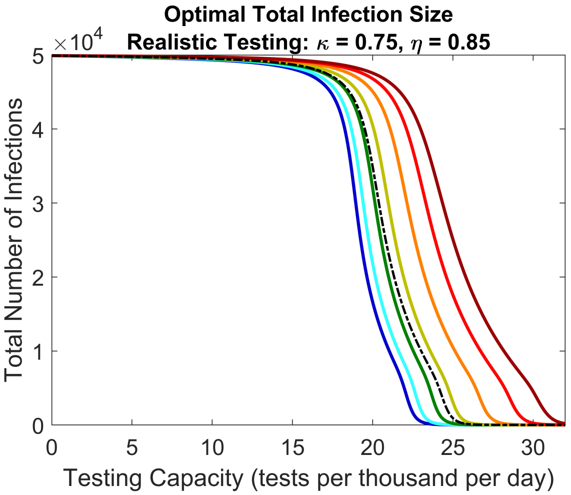


(b) Generic Disease $R_0^{un} = 6.4$: Gamma Distribution Assumptions and Variable Incubation-Latent Offsets

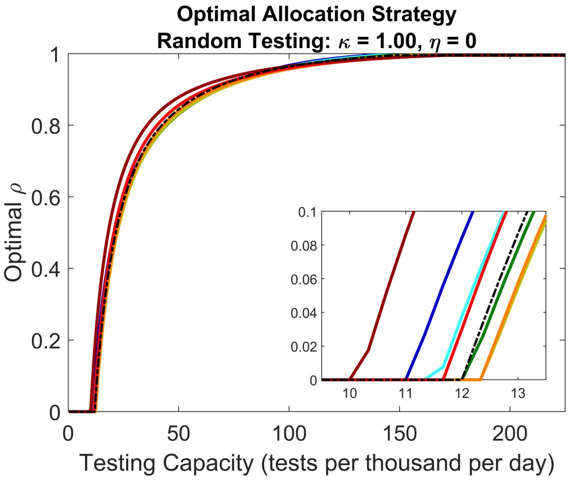


(c) Generic Disease $R_0^{un} = 6.4$: Gamma Distribution Assumptions and Variable Incubation-Latent Offsets

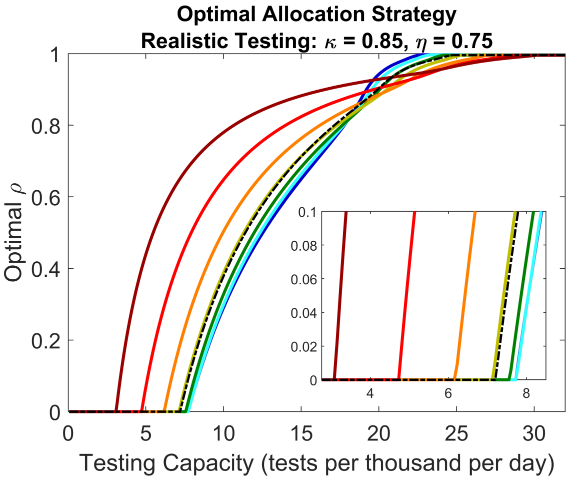


(d) Generic Disease $R_0^{un} = 6.4$: Gamma Distribution Assumptions and Variable Incubation-Latent Offsets

Figure S10: Optimal total infection sizes and corresponding optimal resource allocation strategies for the generic disease with $R_0^{un} = 6.4$, assuming gamma distributions for $f_\varepsilon$ and $f_r$. Curve colors represent different offsets $z = \langle f_I \rangle - \langle f_\varepsilon \rangle$ between mean incubation and latent periods as indicated by the legend in Fig. S10a (measured in units of days). Black dashed curves represent the correlated symptoms assumption where $z$ values are irrelevant. Insets in Figs. S10c and S10d zoom in on strategy threshold testing capacities $C^{th}$ where optimal strategies switch from clinical only to mixed clinical and non-clinical.

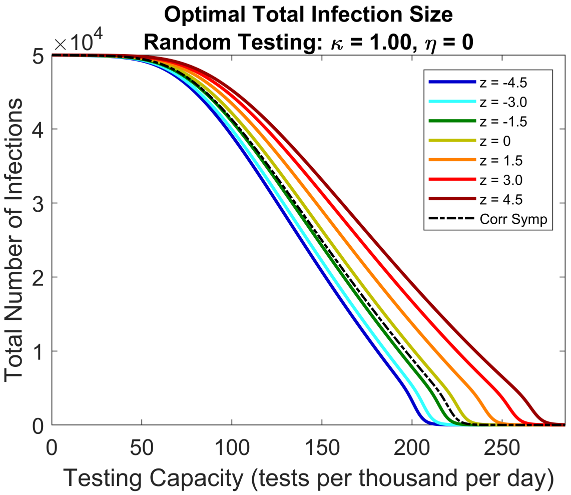


(a) Generic Disease $R_0^{un} = 9.5$: Gamma Distribution Assumptions and Variable Incubation-Latent Offsets

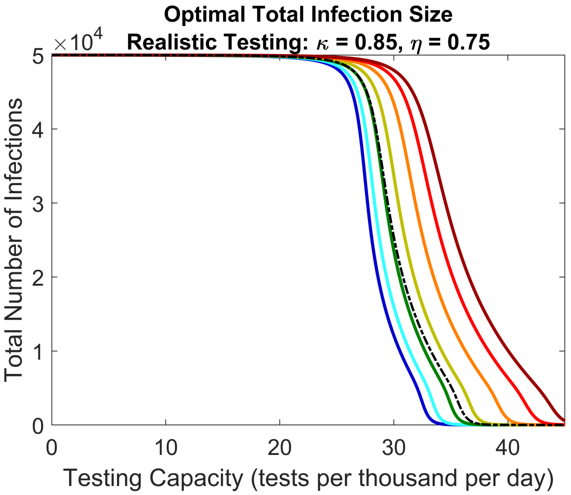


(b) Generic Disease $R_0^{un} = 9.5$: Gamma Distribution Assumptions and Variable Incubation-Latent Offsets

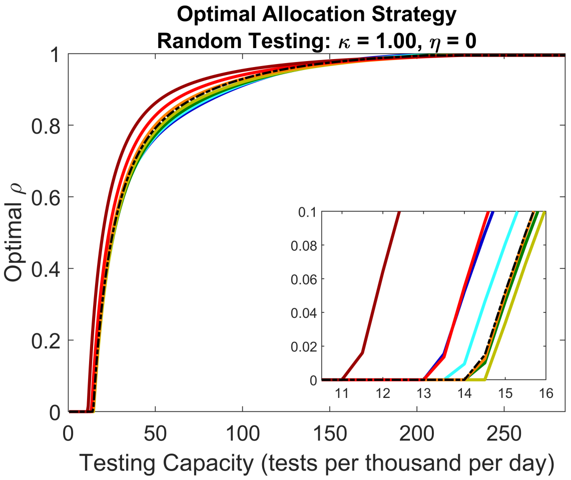


(c) Generic Disease $R_0^{un} = 9.5$: Gamma Distribution Assumptions and Variable Incubation-Latent Offsets

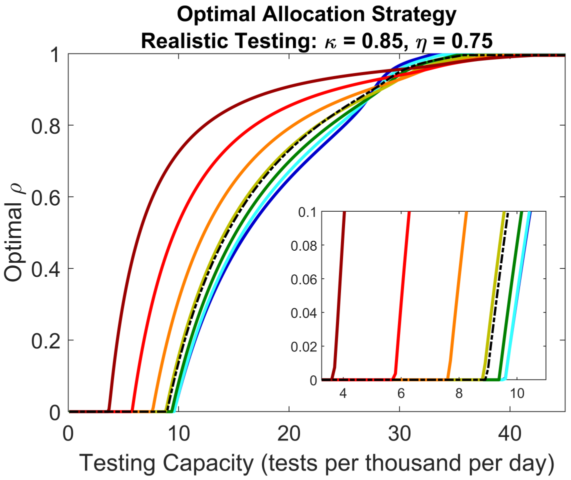


(d) Generic Disease $R_0^{un} = 9.5$: Gamma Distribution Assumptions and Variable Incubation-Latent Offsets

Figure S11: Optimal total infection sizes and corresponding optimal resource allocation strategies for the generic disease with $R_0^{un} = 9.5$, assuming gamma distributions for $f_\varepsilon$ and $f_r$. Curve colors represent different offsets $z = \langle f_I \rangle - \langle f_\varepsilon \rangle$ between mean incubation and latent periods as indicated by the legend in Fig. S11a (measured in units of days). Black dashed curves represent the correlated symptoms assumption where $z$ values are irrelevant. Insets in Figs. S11c and S11d zoom in on strategy threshold testing capacities $C^{th}$ where optimal strategies switch from clinical only to mixed clinical and non-clinical.

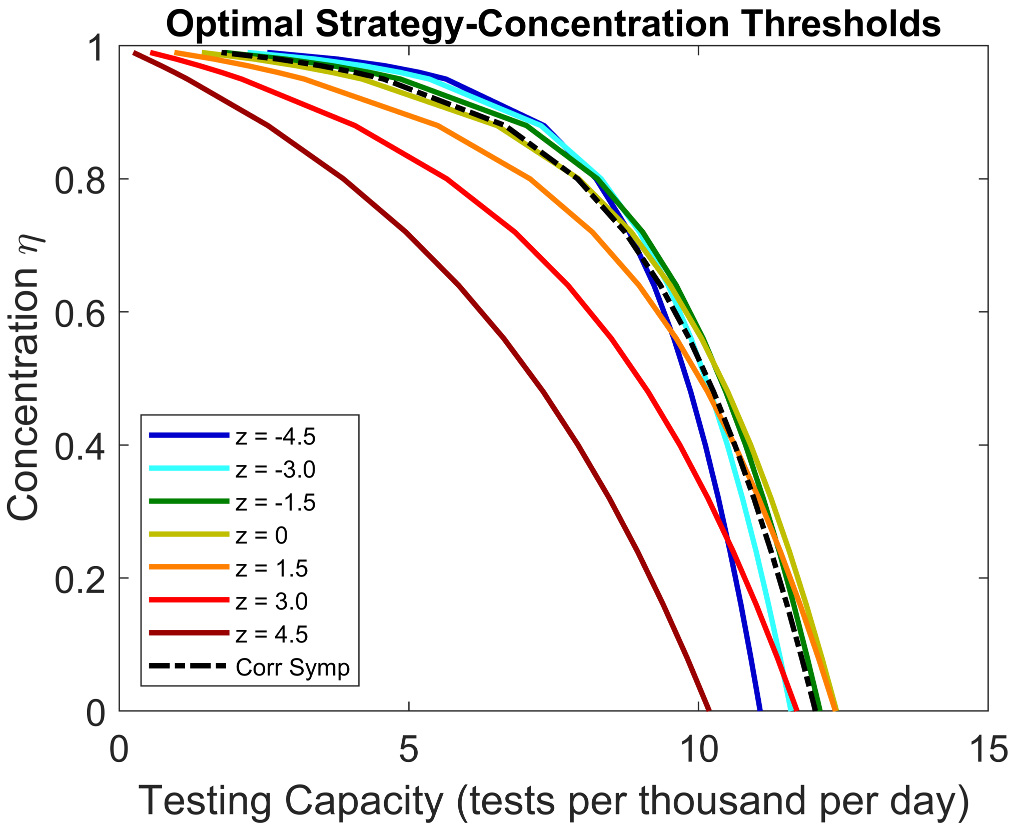


Figure S12: Strategy threshold testing capacities at various concentration levels $\eta$ for the generic disease with $R_0^{un} = 6.4$. Curves represent $(C, \eta)$ values where the strategy threshold capacity $C^{th}$ occurs, assuming either incubation symptoms (colored curves) or correlated symptoms (black dashed curve) with gamma distributed $f_r$ and $f_\varepsilon$. Colors represents different off sets $z$ between the mean incubation and mean latent period, with values in the legend given in units of days. At $(C, \eta)$ points above or to the right of a given curve, optimal strategies are mixed clinical and non-clinical for that $z$ value or symptom assumption. At points below or to the left, optimal strategies are clinical only. The $\eta$ value for which a colored curve crosses the black curve represents a threshold $\eta^{th}$ for which the role of non-clinical testing switches between enhanced and diminished compared to the correlated symptoms assumption.

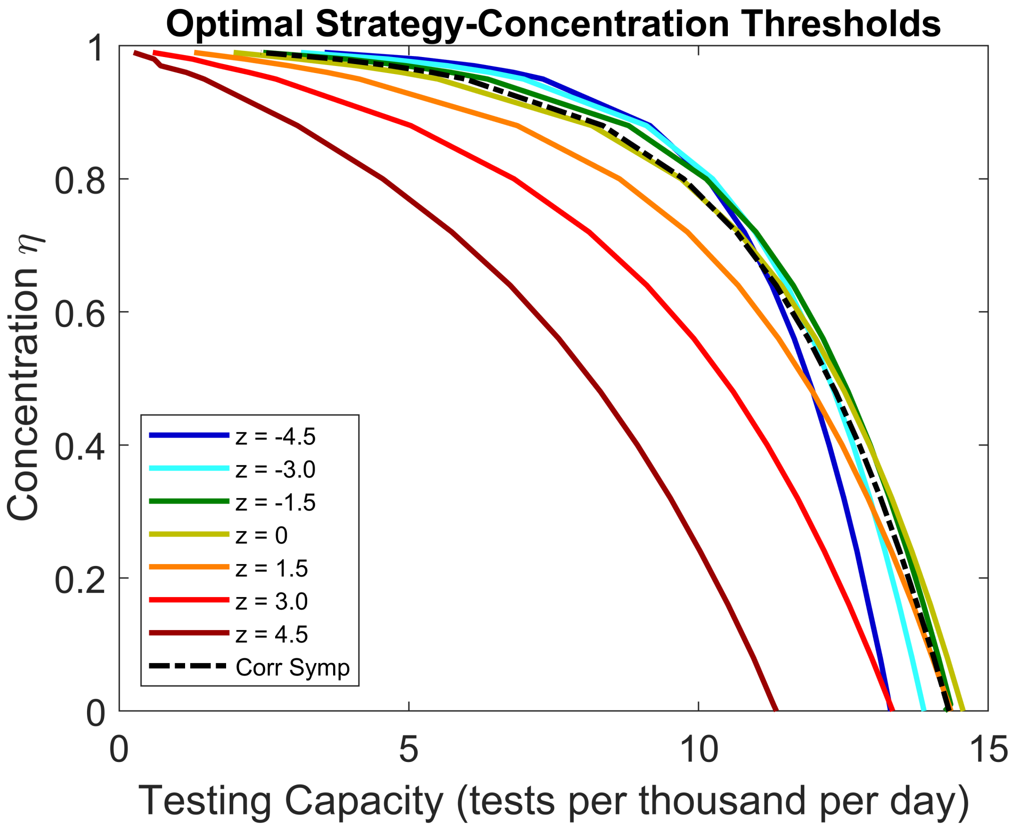


Figure S13: Strategy threshold testing capacities at various concentration levels $\eta$ for the generic disease with $R_0^{un} = 9.5$. Curves represent $(C, \eta)$ values where the strategy threshold capacity $C^{th}$ occurs, assuming either incubation symptoms (colored curves) or correlated symptoms (black dashed curve) with gamma distributed $f_r$ and $f_\varepsilon$. Colors represents different off sets $z$ between the mean incubation and mean latent period, with values in the legend given in units of days. At $(C, \eta)$ points above or to the right of a given curve, optimal strategies are mixed clinical and non-clinical for that $z$ value or symptom assumption. At points below or to the left, optimal strategies are clinical only. The $\eta$ value for which a colored curve crosses the black curve represents a threshold $\eta^{th}$ for which the role of non-clinical testing switches between enhanced and diminished compared to the correlated symptoms assumption.

[2] P. van den Driessche and James Watmough. Reproduction numbers and sub-threshold endemic

equilibria for compartmental models of disease transmission. *Math Biosci*, 180:29–48, 2002. doi: 10.1016/S0025-5564(02)00108-6.
